# Potential impact of catch-up HPV vaccination on HPV prevalence and cervical cancer incidence among women living with HIV in South Africa: results from two mathematical models

**DOI:** 10.64898/2026.01.06.26343544

**Authors:** Carla M Doyle, Minttu M Rönn, Cari van Schalkwyk, Marc Brisson, Nirali Soni, Marie-Claude Boily, Mathieu Maheu-Giroux

**Affiliations:** Department of Epidemiology and Biostatistics, School of Population and Global Health, McGill University, Montréal, QC, Canada; Department of Global Health and Population, Harvard T.H. Chan School of Public Health, Boston, MA, USA; The South African Centre for Epidemiological Modelling and Analysis, Stellenbosch University, Stellenbosch, South Africa; Centre de recherche du CHU de Québec, Québec, Canada; Département de médecine sociale et préventive, Université Laval, Québec, Canada; Centre for Health Informatics, Computing, and Statistics, Lancaster University Medical School, Lancaster, United Kingdom; MRC Centre for Global Infectious Disease Analysis, School of Public Health, Imperial College London, London, UK

**Author notes:** **Address for Correspondence:** 2001 McGill College, Suite 1200, Montréal, QC, Canada H3A 1G1. Contributed equally (Minttu M Rönn, Cari van Schalkwyk). **E-mail addresses of authors:** CMD, MR, CvS, MB, NS, M-CB, MM-G.

**Keywords:** Cervical cancer, HPV, Women living with HIV, HPV vaccination, Mathematical modelling

## Abstract

**Introduction:** Women living with HIV (WLHIV) face an increased risk of cervical cancer (CC). With inequitable HPV vaccine access and a programmatic focus on girls-only school-based delivery, many WLHIV in high HIV prevalence countries remain vulnerable to HPV infection and CC. We assessed the incremental impact of adding catch-up vaccination for WLHIV in South Africa.

**Methods:** We used two independently developed HPV/CC and HIV transmission models to predict the incremental impact of catch-up HPV vaccination for WLHIV compared to routine-only vaccination of girls aged 9-14 (90% cohort coverage) from 2020 onward using a nonavalent vaccine (lifelong 100% protection). We assessed two catch-up scenarios vaccinating WLHIV aged 15-24 or 15+ (attaining 90% cohort coverage for at least three years), maintaining baseline CC screening. We report the predicted median annual prevalence of vaccine-type high-risk HPV (VT HR-HPV), CC incidence, and cumulative fraction of CC averted compared to routine-only vaccination among WLHIV overall (age-standardized) and by age.

**Results:** With routine-only vaccination, overall coverage among WLHIV remained substantially (>50%) lower than in all women for 35-40 years, with gaps persisting even after 80 years. Adding catch-up vaccination for WLHIV aged 15-24 increased coverage and benefits mainly among young WLHIV, with the largest annual reductions (relative to routine-only vaccination) in VT HR-HPV prevalence and CC incidence for WLHIV <30 years, reaching up to 36-52% across models within 15 years and 38-100% after 15-25 years, respectively. If catch-up included WLHIV aged 15+, overall vaccination coverage among WLHIV would immediately increase to 90%, extending benefits to older WLHIV –reducing peak annual relative reductions in CC incidence among WLHIV 50+ by an extra 19%-points compared to catch-up vaccination of WLHIV aged 15-24, and shifting the peak 25 years earlier. Over 55-60 years, catch-up vaccination of WLHIV aged 15-24 and 15+ could avert up to 3-8% and 14% of CC cases among WLHIV, respectively.

**Conclusions:** Catch-up vaccination of WLHIV can reduce their CC burden in the short to medium term, even in the presence of girls-only routine vaccination programs with high cohort coverage. To maximize impact, vaccines should be offered to WLHIV of all ages, not only younger women.

## Introduction

Cervical cancer (CC) is preventable and curable when detected early. Yet, in 2022, approximately 660,000 women were diagnosed with, and 350,000 died from CC globally^1^. The CC burden is disproportionately high in certain regions and populations, largely due to inequitable prevention and treatment service access and the prevalence of risk factors, including HIV^2–4^. Eastern, southern, central, and west Africa –home 26 million people living with HIV (PLHIV)^5^–have the highest CC incidence and mortality^1, 3^. The World Health Organization (WHO) set programmatic CC elimination goals^6^; however, high HIV prevalence settings may require additional strategies.

Women living with HIV (WLHIV) face increased human papillomavirus (HPV) infection vulnerabilities, including heightened risks of acquiring and experiencing persistent high-risk HPV (HR-HPV) infection, and faster progression to cervical lesions and CC^7, 8^. While HIV antiretroviral therapy (ART) partially restores immune function, WLHIV on ART still remain at elevated risk for these HPV-related morbidities^7^. HPV vaccines are safe and produce a robust immunological response in PLHIV^9, 10^, and can reduce WLHIV’s CC risk and burden.

Over the past 20 years, three types of HPV vaccines emerged: bivalent (protecting against HPV16/18), quadrivalent (HPV6/11/16/18), and nonavalent (HPV6/11/16/18/31/33/45/52/58)^11^. Initially licenced under 3-dose schedules, these vaccines showed near 100% seropositivity and efficacy against HPV-related disease outcomes, with 2- and, recently, 1-dose schedules showing non-inferior immunogenicity and comparable protection against persistent HPV infection^11, 12^. Despite these advancements, vaccine access across Africa remains suboptimal^13^. Rwanda launched Africa’s first national HPV immunisation program in 2011^13, 14^; by 2023, only 54% of countries had established programs, and uptake varies (first-dose coverage from 5-99%)^13^. Most of these programs focused on girls-only, school-based delivery of cohorts typically in early adolescence^13, 15, 16^, leaving many older women, and particularly WLHIV, vulnerable to HPV acquisition and CC.

South Africa’s HIV prevalence is among the highest, with 9% of adolescent girls and young women aged 15-24 years living with HIV (AGYW-LHIV) in 2024^17^. The country’s public CC control strategy includes cytology-based screening and, since 2014, school-based 2-dose bivalent vaccination of girls generally aged 9-12 years^18^. National guidelines recommend screening 30-50-year-old women three times (every 10 years), or upon HIV diagnosis and every three years for WLHIV (or annually after a positive screen)^18^. A previous mathematical model comparison study assessed vaccine and screening strategies for CC elimination by 2120 in South Africa^19^. While it found that vaccination and twice-lifetime screening could achieve elimination in all women, it did not fully examine the added benefits of catch-up vaccination for WLHIV.

We explored the incremental impact –beyond routine vaccination of young girls– of catch-up vaccination for WLHIV in South Africa on HPV and CC outcomes using two mathematical models. We investigated how benefits accrue over time and across age groups, and considered variations in vaccination age, type, coverage, and duration of immunity, and ART coverage (e.g., following global HIV funding cuts^20^).

## Methods

### Model overviews

We leveraged two independently developed mathematical models of heterosexual HPV-HIV co-transmission in South Africa: *Det_HPV-HIV*^19^ and *MicroCOSM-HPV*^19, 21–23^. Both models simulate HIV and 13 HR-HPV genotypes (vaccine types 16/18/31/33/45/52/58 and non-vaccine types 35/39/51/56/59/68) and HIV/HPV interactions that increase the risk of persistent HPV and associated disease progression in WLHIV according to ART status (ART partially decreases persistent HPV risk and slows CC progression in WLHIV^24, 25^)*. Det_HPV-HIV* further assumes that PLHIV have an increased HPV acquisition risk, and vice versa. CC screening is incorporated to mimic the implementation in South Africa. HPV vaccination is modelled from 2014 (*MicroCOSM-HPV*) or 2020 (*Det_HPV-HIV*), with scenario-specific coverage and eligibility (Table 1). Finally, both models were calibrated under Bayesian frameworks to South African demographic and epidemiologic data up to 2020.

**Table 1.**
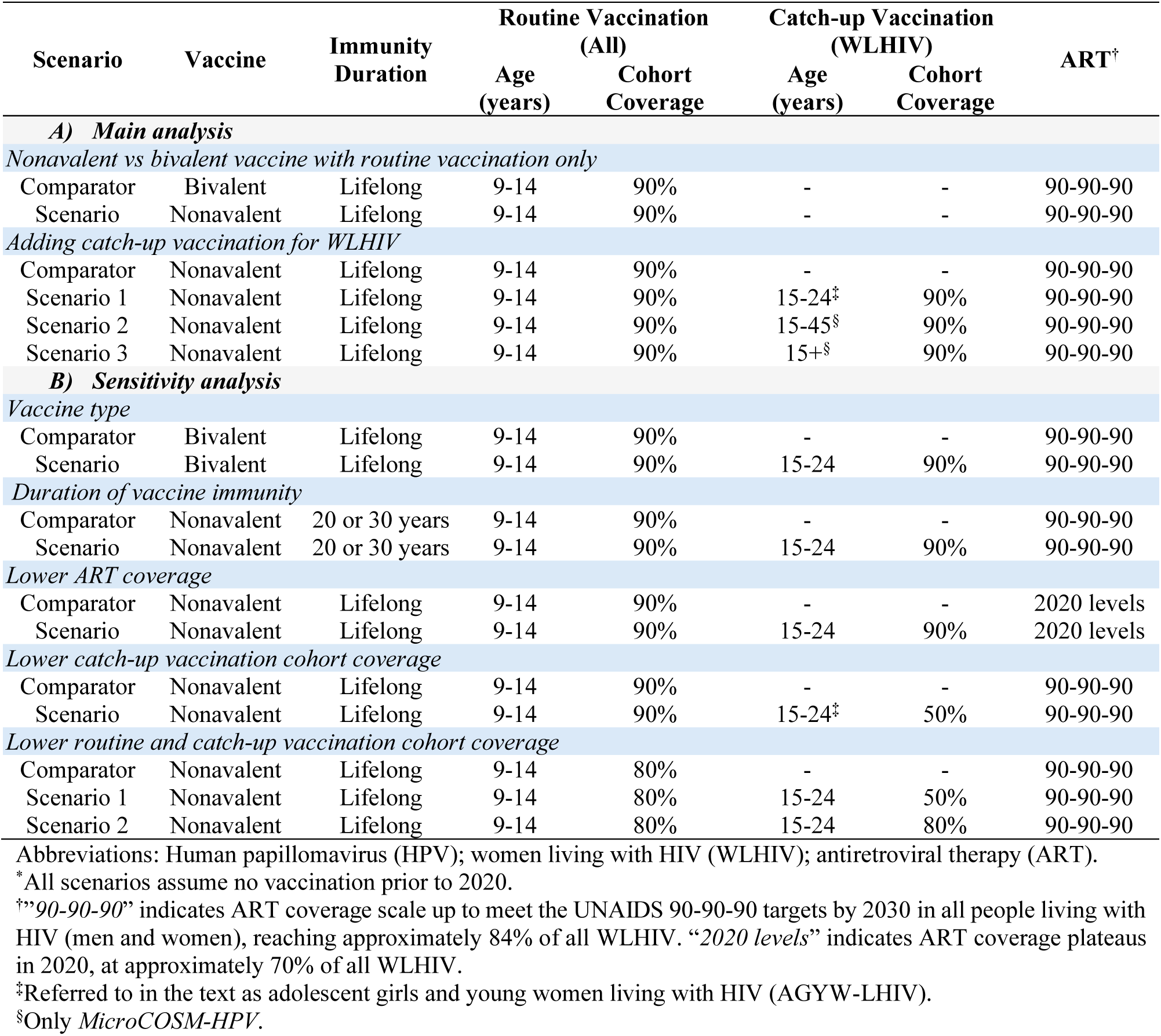
Modelled HPV vaccination scenarios in A) main analysis and B) sensitivity analysis*.

### Det_HPV-HIV: compartmental model

*Det_HPV-HIV*^19^ is a dynamic compartmental model simulating three groups of HR-HPV (16/18, 31/33/45/52/58, and others not in the nonavalent vaccine) from 1950 onward, with HIV transmission introduced in 1985. The model is stratified by sex, 5-year age groups (from 9-14 to 70-74 years), and sexual-activity levels (low/medium/high). Condoms and male circumcision for HIV prevention are modelled, with time-varying coverage per sex act. ART is introduced in 2004 (when South Africa’s publicly-funded program launched), with coverage increasing towards 70% of PLHIV by 2020. Between 2000-2012, CC screening was offered in South Africa, but coverage was low^26^. Thus, CC screening and treatment are introduced in 2012, with annual screening uptake increasing to, and plateauing at, 40% in 25-year-olds and 55% in 35- and 45-year-olds in 2017 (without differences by HIV-status)^27^. The model was calibrated to sex- and age-specific data on national demographics, HIV prevalence, ART coverage, HR-HPV and cervical intraepithelial neoplasia grade 2 or worse (CIN2+) prevalence by HIV status, and CC incidence (Figure S1).

### MicroCOSM-HPV: individual-based model

*MicroCOSM-HPV*^19, 21–23^ is an individual-based model simulating each HR-HPV genotype and HIV from 1985 (beginning with 1 million individuals), considering sex, continuous age, and sexual-risk levels (low/high). The protective effect of condoms against HIV and HPV is considered, with coverage varying by time, sex, age, and relationship type. ART is modelled from 2000 onward, with uptake varying by time, sex, and disease stage, reaching 70% coverage among PLHIV by 2020. In 2000, CC screening and treatment are introduced, with screening probabilities depending on age, time since last screen, and ART-status among WLHIV estimated in calibration. Modelled screening coverage in 2019 was 45% among women not living with HIV aged 30-60 and 33% among WLHIV aged 25-60. The model was calibrated to national HIV prevalence, type-specific HPV prevalence (by age and sex), CC screening coverages, CIN1 and CIN2/3 prevalence, and CC incidence (Figure S1).

### Vaccination scenarios

Each model simulated scenarios of routine vaccination for girls. Additional scenarios added on catch-up vaccination for WLHIV in 2020 (Table 1), implemented to reach and maintain a target cohort coverage for three years in *Det_HPV-HIV* and continuously in *MicroCOSM-HPV*. Unless otherwise specified, scenarios attain 90% cohort coverages (i.e., 90% of age-eligible girls or WLHIV are vaccinated), are based on the nonavalent vaccine, assume 100% lifelong vaccine protection (without cross-protection against non-vaccine types), and maintain CC screening assumptions and coverage from 2019 onward. ART scale-up is modelled to meet 81% coverage in all PLHIV by 2030 (consistent with 90-90-90 targets), reaching approximately 84% among WLHIV in both models (Figure S2). As HIV diagnosis is not modelled explicitly, catch-up vaccination is applied to all WLHIV irrespective of diagnosis status.

We first estimated the impact of using the nonavalent vaccine (rather than bivalent) from 2020 onward for routine-only vaccination (no catch-up) to understand the effect of the highest-valency vaccine available. Compared to routine-only vaccination, we then assessed the incremental impact of adding catch-up vaccination among AGYW-LHIV and, using *MicroCOSM-HPV*, among WLHIV aged 15-45 or 15+.

In sensitivity analyses, we assessed the incremental impact of catch-up vaccination among AGYW-LHIV: 1) using the bivalent vaccine; 2) reducing the duration of vaccine-induced immunity (20/30 years); 3) lowering ART coverage (assuming coverage among WLHIV plateaued near 70% in 2020; Figure S2); and 4) lowering cohort vaccination coverages for routine and catch-up vaccination (Table 1).

### Outcomes

We predicted annual HPV vaccination coverage, annual VT and all HR-HPV prevalence, and annual and cumulative number of new CC cases for each scenario (2019-2100). We measured the additional population-level impact of each catch-up scenario compared to the relevant comparator scenario (see Table 1) over 2020-2100 by the annual relative reduction in HR-HPV prevalence, the annual relative reduction in CC incidence rates, and the cumulative fraction of CC cases averted. Each outcome was calculated among WLHIV overall (age-standardized by 5-year age groups to the South African UN Population Projections^28^) and stratified by age.

### Ethics

This study used simulated outputs from previously published mathematical models and did not require ethical approvals.

## Results

### Vaccination coverage across scenarios

Following routine-only vaccination, overall coverage among WLHIV steadily increased, as each successive cohort of girls was vaccinated (Figure 1A). However, given the increasingly older age profile of WLHIV due to declines in HIV incidence (Figure S3A-B), their vaccine coverage consistently lagged that of all women, remaining >50% below for 35-40 years (Figures 1B, S4). Adding catch-up vaccination of AGYW-LHIV increased overall coverage among WLHIV 7-9-fold in 2020 and more than 2-fold until 2040 compared to routine-only vaccination (with limited impact on coverage in all women), but the coverage gap between WLHIV and all women still persisted for nearly 80 years (Figures 1B, S4). Eliminating this disparity required expanding catch-up to WLHIV aged 15-45 or 15+, which brought overall coverage among WLHIV to 63% and 90% in 2020, respectively, and increased coverage among all women 2- to 3-fold compared to routine-only vaccination (Figures 1A-B, S4). Furthermore, while coverage among AGYW-LHIV rose rapidly across all catch-up scenarios, without including older WLHIV (15-45 or 15+ years), it took 65-75 years to reach high coverage (>70%) among WLHIV aged 50+, who are most likely to develop CC (Figure S5).

**Figure 1.**
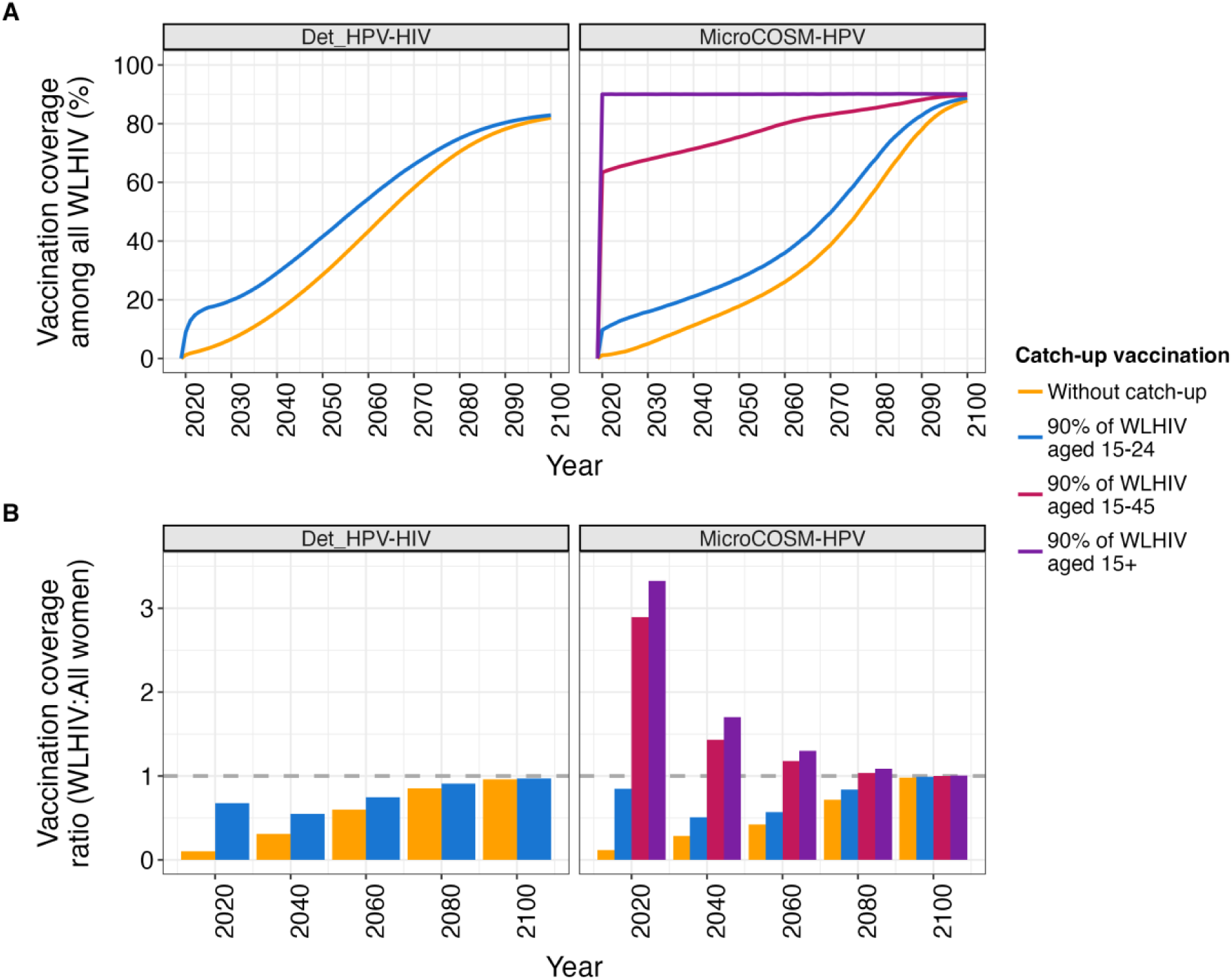
Overall vaccination coverage among women living with HIV (WLHIV). <u>Panel A</u>: The modelled overall vaccination coverage among all WLHIV over 2019-2100 under scenarios of routine vaccination in girls aged 9-14 years (90% cohort coverage) without and with catch-up vaccination for WLHIV aged 15-24 (AGYW-LHIV), WLHIV aged 15-45, and WLHIV aged 15+ (90% cohort coverage). <u>Panel B</u>: The ratio of overall vaccination coverage between WLHIV and all women at 20-year intervals between 2020-2100 under those same scenarios. Each panel presents the median estimates per model. The colour indicates the scenario.

### Impact of routine-only vaccination

The median baseline (2019) VT HR-HPV prevalence among WLHIV was 27% in both models (Figure S6A), with important age differences (Figure S8). Without vaccination, the prevalence among WLHIV modestly declined until 2030 (alongside ART scale-up), and stabilized or continued to gradually decrease to 18-23% over 2030-2100 (Figures S6A, S8). Routine-only vaccination with the bivalent vaccine halved baseline VT HR-HPV prevalence among WLHIV after approximately 60-75 years (by 2078-2093), and declined to 12-13% across models in 2100 (Figure S7A), mainly driven by greater and more sustained decreases in younger women with high vaccination coverage (Figure S8). Routine-only vaccination with the nonavalent vaccine halved the baseline prevalence among WLHIV about 40-50 years sooner than the bivalent vaccine (within 20 years, by 2038-2041), and surpassed a 90% decline after nearly 65 years (Figure S7A). Furthermore, all age groups experienced substantial prevalence declines with the nonavalent vaccine (Figure S8): VT HR-HPV prevalence was <5% within 11-36 years among <30-year-olds and 30-65 years among those aged 30+, with overall prevalence reaching <2% by 2092-2093 (Figure S7A). Figures S6B and S7B present all HR-HPV type prevalence.

The median baseline CC incidence rate among WLHIV was 91-102 cases per 100,000 across models (Figure S5C), with higher rates in older WLHIV (108-264 per 100,000 cases in those aged 30+ across models; Figure S9). Like HR-HPV prevalence, without vaccination CC incidence among WLHIV declined until 2030-2050 (Figure S6C), especially in those aged 40+ (Figure S9). However, despite the long-term HR-HPV prevalence trends and subsequently stable age-stratified CC incidence estimates, when age-standardized, CC incidence in all WLHIV was predicted to increase to 104-107 cases per 100,000 by 2100 (Figure S6C). Routine-only vaccination halved CC incidence among WLHIV after 52-72 years (by 2072-2092) and 48-52 years (by 2068-2072) with the bivalent and nonavalent vaccine, respectively, reaching 38-44 and 24-26 cases per 100,000 by 2100 (Figure S7C). All age groups experienced accelerated declines in CC incidence under routine-only vaccination –in the short-term among WLHIV <40 years and over time in WLHIV 40+– that were only slightly more pronounced with the nonavalent vaccine (Figure S9).

Compared to all women, who showed similar long-term HR-HPV prevalence trends and stable CC incidence without vaccination (Figure S6A-C), rates were consistently higher for WLHIV, even after routine-only vaccination (Figure S7A-C). However, baseline estimates among all women were halved by routine-only vaccination (bivalent or nonavalent) at comparable times as WLHIV. Routine-only vaccination with the nonavalent vaccine had a greater impact on HR-HPV prevalence than CC incidence, among both WLHIV and all women (Figure S10A-C). This translated into near negligible added CC benefits by using the nonavalent over the bivalent vaccine for routine-only vaccination in the short- to mid-term, averting <2% of cumulative CCs among WLHIV and all women within 28-55 years (Figure S11A-B). These added benefits increased gradually over time, but less substantially for WLHIV than all women, preventing 4-14% and 6-20% of CCs among WLHIV and all women, respectively, after 80 years (Figure S11A-B).

### Impact of adding catch-up vaccination for WLHIV on HPV prevalence

Alongside increased vaccination coverage, adding catch-up vaccination of AGYW-LHIV and WLHIV 15+ accelerated VT HR-HPV prevalence declines among WLHIV–achieving 50% and 90% declines from baseline up to 6 years earlier than routine-only vaccination across scenarios in both models (Figure 2A; see Figure S12 for all HR-HPV types). Compared to routine-only vaccination, catch-up vaccination of AGYW-LHIV or WLHIV 15+ (*MicroCOSM-HPV* only) resulted in annual relative reductions in VT HR-HPV prevalence among WLHIV by up to 12-13% or 22%, respectively, within five years and 12-25% or 37% within 40 years across models (Figure 2B).

**Figure 2.**
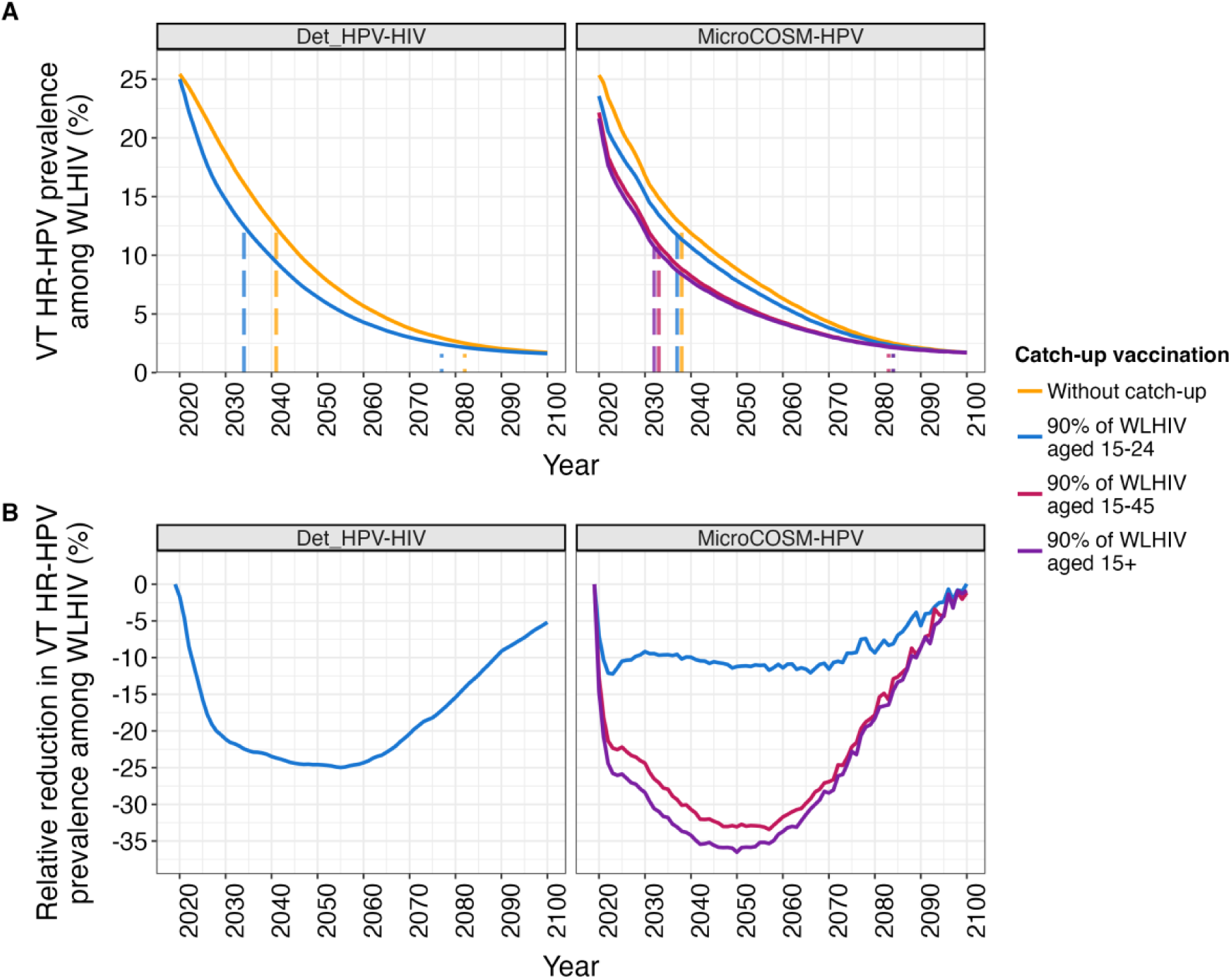
Impact of catch-up vaccination on vaccine type (VT) high-risk HPV (HR-HPV) prevalence among women living with HIV (WLHIV). The annual estimated, age-standardized VT HR-HPV prevalence among WLHIV over 2019-2100 under scenarios of routine vaccination in girls aged 9-14 years (90% cohort coverage) without and with catch-up vaccination for WLHIV aged 15-24 (AGYW-LHIV), WLHIV aged 15-45, and WLHIV aged 15+ (90% cohort coverage). <u>Panel A</u>: absolute prevalence estimates. The vertical dashed and dotted lines indicate the year when VT HR-HPV prevalence declined by 50% and 90% compared to the baseline (2019), respectively. <u>Panel B</u>: relative reduction in prevalence of different catch-up vaccination scenarios compared to routine-only vaccination. Each panel presents the median estimates per model. The colour indicates the scenario.

Following catch-up vaccination for AGYW-LHIV, annual relative reductions (compared to routine-only vaccination) in VT HR-HPV prevalence among <30-year-old WLHIV reached up to 36-52% across models within 15 years (2022-2036), while attaining <36% across age groups of WLHIV 30+ in both models after 15 years or more (Figure 3A). Expanding catch-up to older WLHIV substantially benefited them (Figures 3B, S13-16). When vaccinating WLHIV 15+ years, the maximum annual relative reductions (compared to routine-only vaccination) in VT HR-HPV prevalence occurred at the same time (2035-2045 for 30-49 year-olds) or 28 years sooner (2055 for 50+-year-olds), and were further increased by 10-13%-points and 29%-points in 30-49 and 50+-year-olds, respectively, than when only vaccinating AGYW-LHIV (Figures 3B, S13-14).

**Figure 3.**
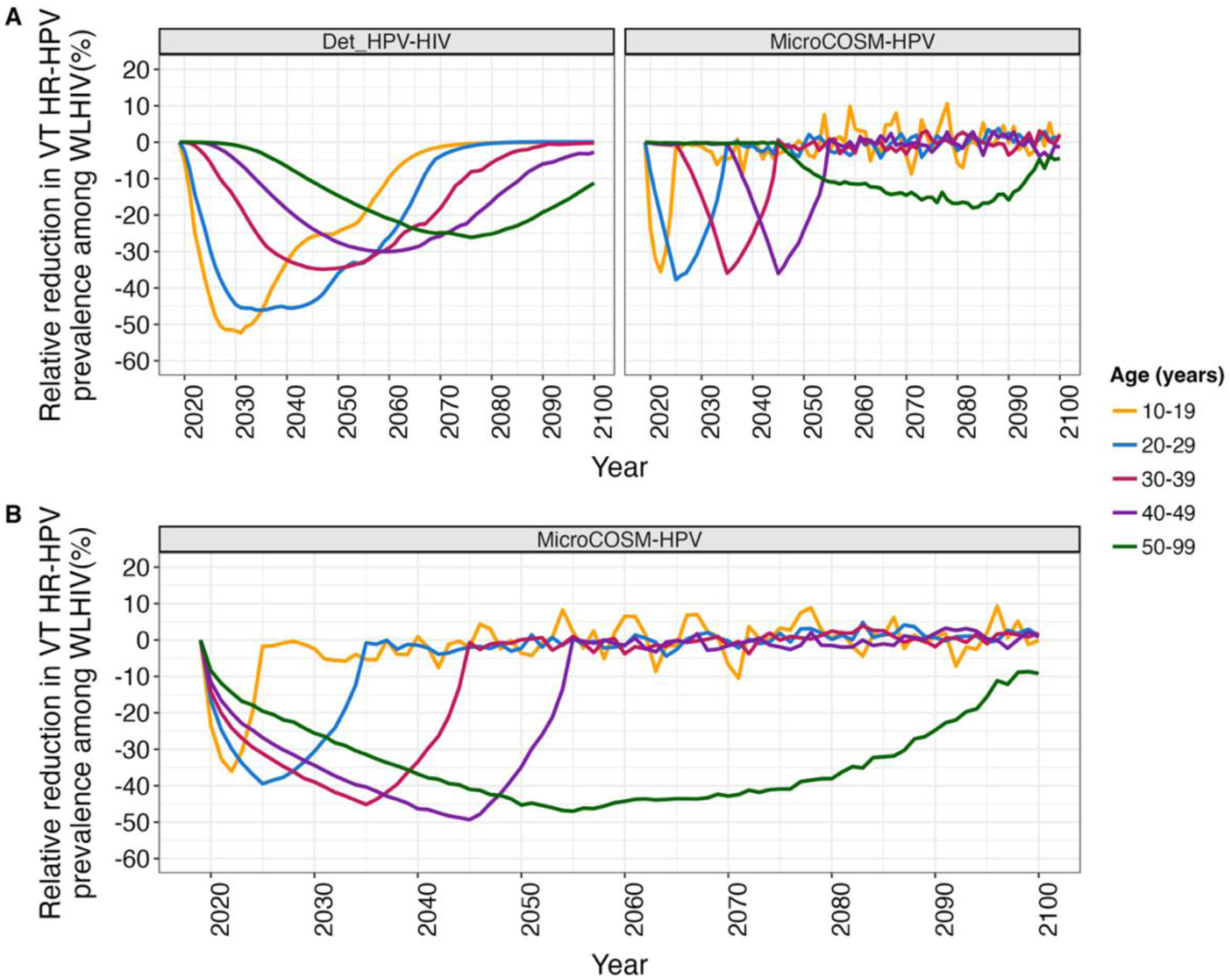
Age-stratified relative reductions in vaccine type (VT) high-risk HPV (HR-HPV) prevalence among women living with HIV (WLHIV) compared to routine-only vaccination. The relative reduction in annual estimated, age-stratified VT HR-HPV prevalence among WLHIV over 2019-2100 under scenarios of routine vaccination in girls aged 9-14 years (90% cohort coverage) with catch-up vaccination for WLHIV aged 15-24 (AGYW-LHIV; Panel A) and WLHIV aged 15+ (Panel B) compared to routine-only vaccination (90% cohort coverage). Each panel presents the median estimates per model. The colour indicates the age category.

### Impact of adding catch-up vaccination for WLHIV on CC incidence

Catch-up vaccination for AGYW-LHIV and WLHIV 15+ accelerated declines in CC incidence, achieving a 50% decline compared to baseline 4-7 (by 2064-2065 across models) and 18 years earlier (*MicroCOSM-HPV*) than routine-only vaccination, respectively (Figure 4A). Compared to routine-only vaccination, catch-up of AGYW-LHIV began reducing annual CC incidence (relative reduction >5%) within 13-25 years across models (by 2033-2045), with reductions peaking at 14% and 21% after more than 55 years (between 2076-2091; Figure 4B). Expanding catch-up to WLHIV 15+ advanced the impacts on CC incidence, with annual relative reductions (compared to routine-only vaccination) >5% 12 years sooner than catch-up of AGYW-LHIV, and increasing to 35% within 30 years (by 2060; Figure 4B).

**Figure 4.**
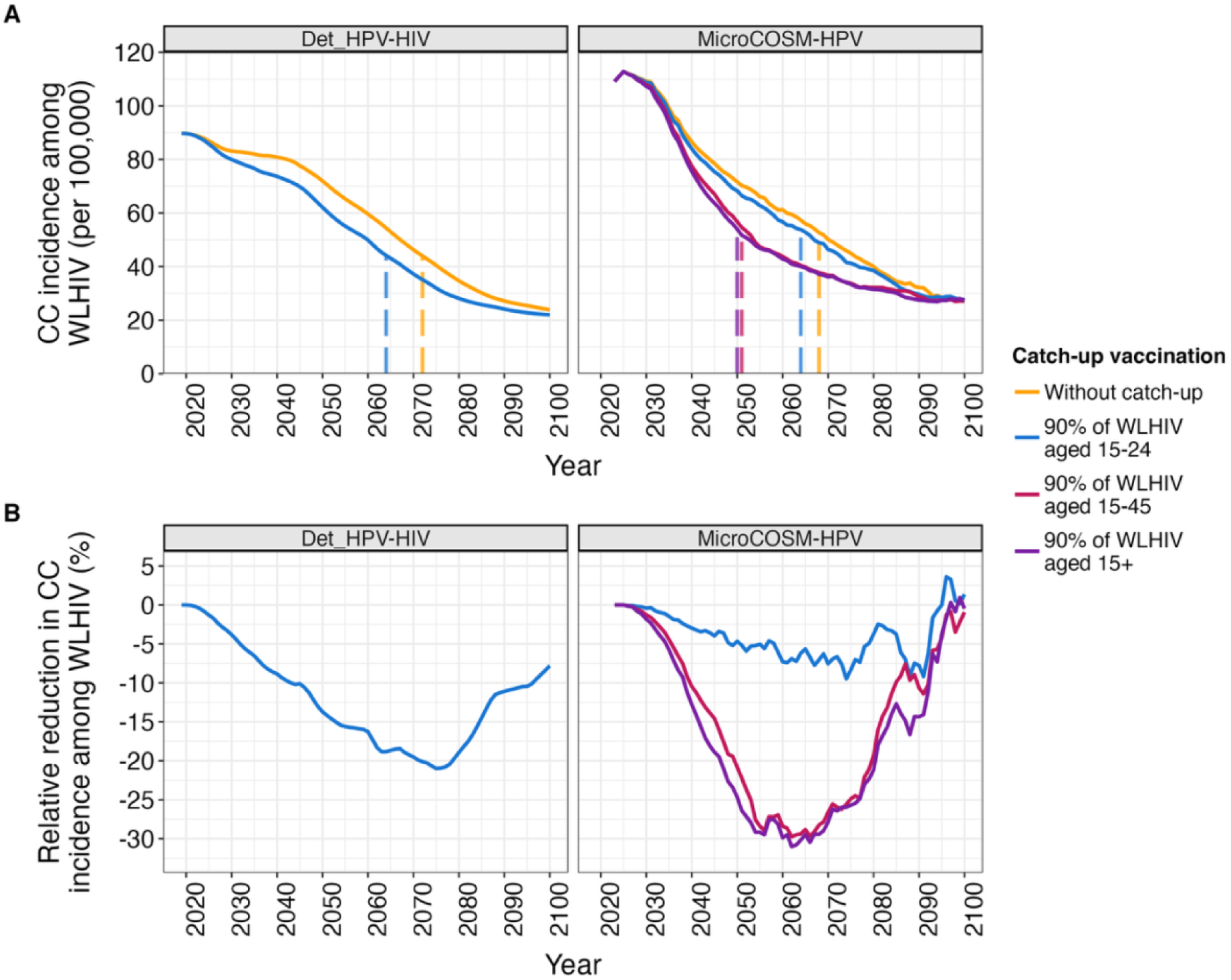
Impact of catch-up vaccination on cervical cancer (CC) incidence among women living with HIV (WLHIV). The annual estimated, age-standardized CC incidence among WLHIV over 2019-2100 under scenarios of routine vaccination in girls aged 9-14 years (90% cohort coverage) without and with catch-up vaccination for WLHIV aged 15-24 (AGYW-LHIV), WLHIV aged 15-45, and WLHIV aged 15+ (90% cohort coverage). <u>Panel A</u>: absolute CC incidence estimates. The vertical dashed lines indicate the year when CC incidence declined by 50% compared to the baseline (2019). <u>Panel B</u>: relative reduction in CC incidence of different catch-up vaccination scenarios compared to routine-only vaccination. Each panel presents the median estimates per model. The colour indicates the scenario. A 5-year simple moving average was applied to the *MicroCOSM-HPV* estimates to smooth stochastic variations.

Following catch-up vaccination of AGYW-LHIV, annual relative reductions in CC incidence reached up to 38-100% among <30-year-old WLHIV across models after approximately 15-25 years (between 2034-2053), while reaching a maximum of 21-36% across age groups of WLHIV 30+ after 25 years or more (between 2043–2091) across models (Figures S17-S18). Like HR-HPV prevalence, benefits to WLHIV 30+ began earlier and were higher when catch-up included older WLHIV. When vaccinating WLHIV 15+ years, the maximum annual relative reductions (compared to routine-only vaccination) in CC incidence further increased by 3%-points around the same time (2056-2048) or 19%-points 25 years sooner (2066) in 30-49 and 50+ year-olds, respectively, than when only vaccinating AGYW-LHIV (*MicroCOSM-HPV*, Figures S17-18).

### Cumulative CC averted by adding catch-up vaccination for WLHIV

Catch-up vaccination of AGYW-LHIV gradually averted more CC cases among WLHIV than routine-only vaccination, taking 15-33 years to avert an extra 2% of cumulative CC cases (by 2035-2053) and 55 years to avert an extra 3-8% across models (by 2074 in both), with little additional benefits thereafter (Figures 5, S19-20). Expanding catch-up to older WLHIV substantially accelerated and increased the additional benefits among WLHIV. Vaccinating WLHIV aged 15+ averted >2% of cumulative CC cases 18 years sooner than catch-up of AGYW-LHIV (by 2035), accumulating benefits for approximately 60 years, averting an extra 14% of CC cases by 2080 (Figures 5, S19).

**Figure 5.**
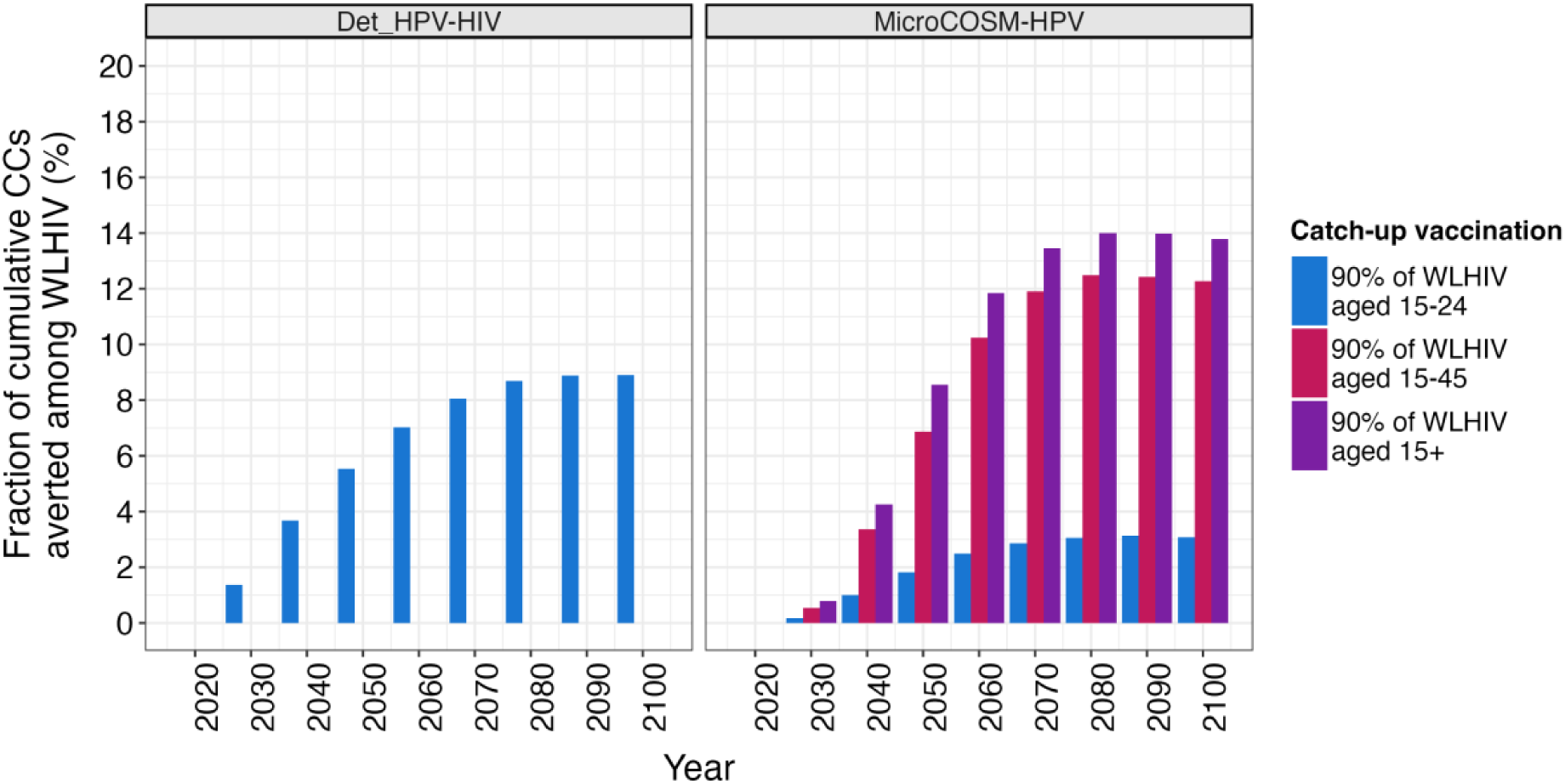
Incremental cumulative fraction of cervical cancers (CC) averted by catch-up vaccination for women living with HIV (WLHIV) compared to routine-only vaccination. The annual estimated, age-standardized cumulative fraction of CCs averted among WLHIV over 2020-2100 (at 10-year intervals) under scenarios of routine vaccination in girls aged 9-14 years (90% cohort coverage) with catch-up vaccination for WLHIV aged 15-24 (AGYW-LHIV), WLHIV aged 15-45, and WLHIV aged 15+ (90% cohort coverage) compared to routine-only vaccination. Each panel presents the median estimates per model. The colour indicates the scenario.

When vaccinating AGYW-LHIV, there is a sequential shift in the timing of the maximum impact across age groups (due to aging of vaccinated cohorts), with cohorts <30 years maximizing within 30-60 years (between 2030-2060), followed by progressively older cohorts until around 2080-2090 (Figures 6A, S21). Vaccinating WLHIV 15+ accelerated impacts and averted a larger fraction of cumulative CC cases compared to routine-only vaccination across all age groups, although the most pronounced increase occurred among WLHIV aged 40-49 and 50+ (Figure 6B, S21). In these age groups, approximately 8% and 21% of cumulative CC cases were averted by 2050 and 2080, respectively, when vaccinating WLHIV 15+ (rather than 3% by 2051 and 4% by 2091, respectively, when vaccinating AGYW-LHIV).

**Figure 6.**
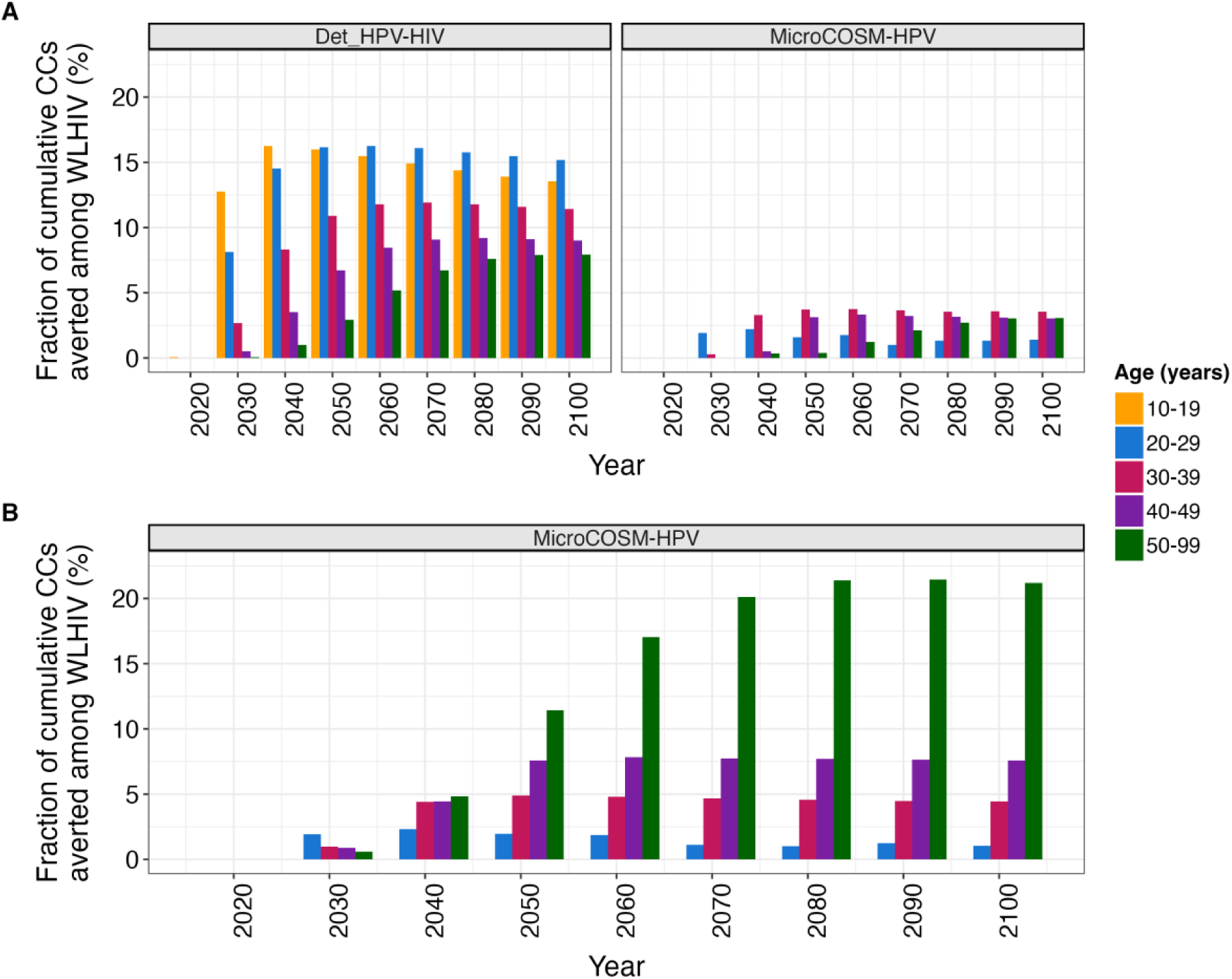
Age-stratified incremental cumulative fraction of cervical cancers (CC) averted by catch-up vaccination for women living with HIV (WLHIV) compared to routine-only vaccination. The annual estimated, age-stratified cumulative fraction of CCs averted among WLHIV over 2020-2100 (at 10-year intervals) under scenarios of routine vaccination in girls aged 9-14 years (90% cohort coverage) with catch-up vaccination for WLHIV aged 15-24 (AGYW-LHIV; <u>Panel A</u>) and WLHIV aged 15+ (<u>Panel B</u>) (90% cohort coverage) compared to routine-only vaccination. Each panel presents the median estimates per model. The colour indicates the age-category. While both models incorporate a low CC incidence for young WLHIV, no CC cases occurred among WLHIV aged 10-19 years in *MicroCOSM-HPV* in our set of stochastic simulations.

### Sensitivity analyses

The incremental impact of catch-up vaccination for AGYW-LHIV (90% cohort coverage) on CC cases compared to routine-only vaccination would be reduced from 2040 onward if vaccine immunity lasts 20/30 years instead lifelong or if the bivalent vaccine is used (Figure 7). When routine and catch-up vaccination coverage were both 80%, results were similar to our main analysis; however, reducing catch-up cohort coverage for AGYW-LHIV to 50% attenuated the incremental impact, being 45% lower than when cohort coverage is 90% by 2080 (Figure 7). Applying catch-up vaccination to WLHIV on ART (*MicroCOSM-HPV*) more gradually increased vaccine coverage among all WLHIV, delaying and attenuating impacts on CC; by 2080, the cumulative fraction of CC cases averted by catch-up for AGYW-LHIV and WLHIV 15+ (compared to routine-only vaccination) would be 30% and 22% lower, respectively, than when vaccinating all WLHIV (Figure S22). Finally, the impact of catch-up vaccination of AGYW-LHIV (90% cohort coverage) under constant ART coverage from 2020 influenced the results differently across models (slightly lower in *Det_HPV-HIV* and higher in *MicroCOSM-HPV*), consistent with differences in duration of catch-up vaccination and HIV and ART interactions with HPV and CC across models (Figure 7).

**Figure 7.**
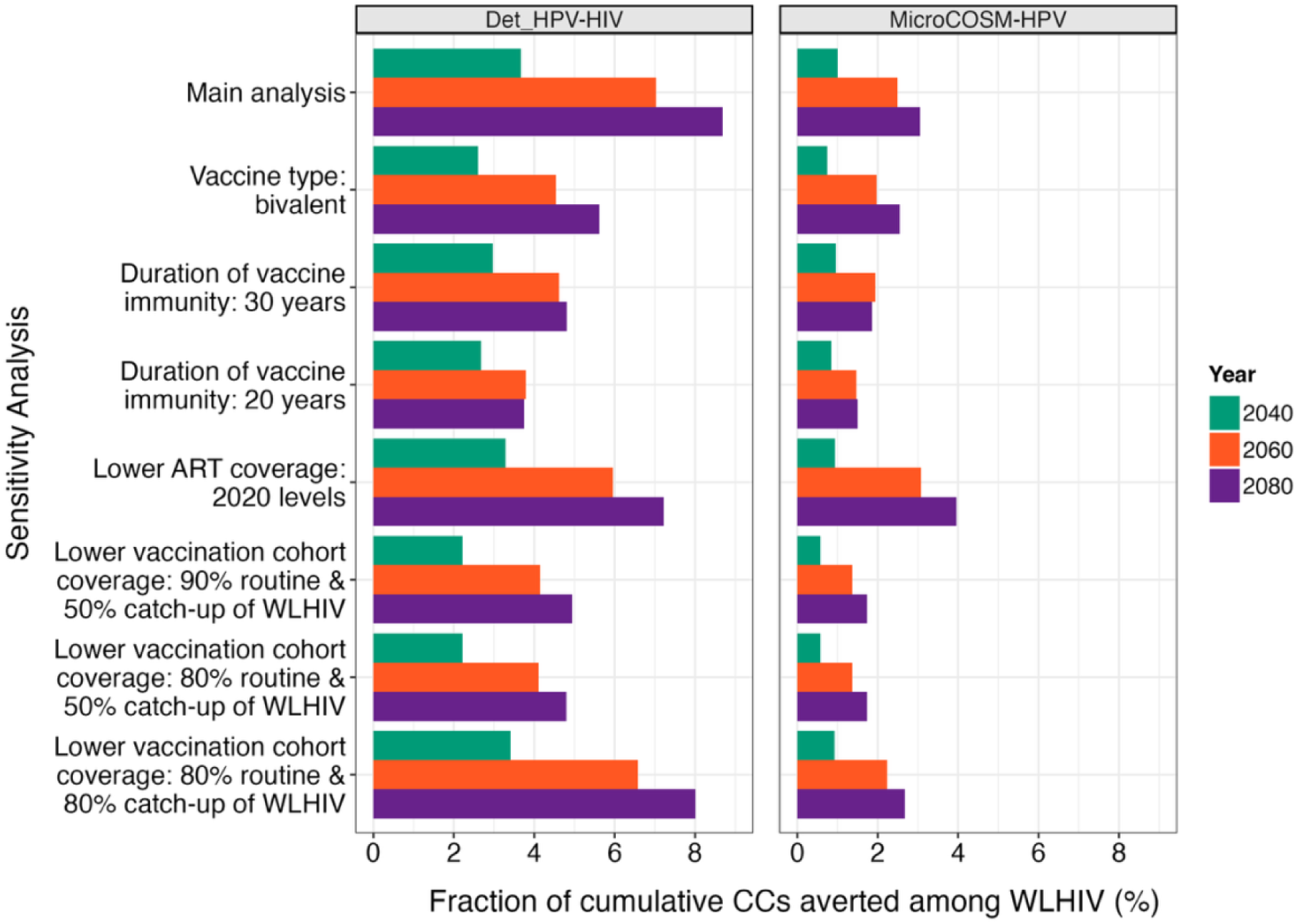
Sensitivity analysis of incremental impact of catch-up vaccination of WLHIV (vs routine-only vaccination) on cumulative fraction of cervical cancers (CC) averted among WLHIV between 2020 and 2040, 2060, and 2080 under alternative conditions. The main analysis presented assumes catch-up vaccination for WLHIV aged 15-24 years (AGYW-LHIV) compared to routine-only vaccination of girls aged 9-14 years (both with 90% cohort coverage) with the nonavalent vaccine, lifetime vaccine immunity, and ART scaling up until 2030 to approximately 84%. Sensitivity analyses explored the same scenarios, but using the bivalent vaccine (lifelong immunity), shorter immunity duration (20 and 30 years; nonavalent vaccine), lower ART coverage (plateauing in 2020 near 70%), lower catch-up cohort coverage (50%), and lower routine (80%) and catch-up vaccination cohort coverage (50% and 80%) (details in Table 1B). Each panel presents the median estimates per model. The colour indicates the year.

## Discussion

Routine HPV vaccination of young girls is fundamental for CC prevention and will importantly reduce HPV and CC in South Africa. Our study used two dynamic transmission models to further investigate the impact of vaccinating WLHIV in this high HIV prevalence setting. We found that adding catch-up vaccination for WLHIV aged 15+ could avert up to 14% more CC cases than routine-only vaccination over 55-60 years. Restricting catch-up to AGYW-LHIV would avert a smaller fraction of all CC cases (3-8%). As WLHIV in South Africa are generally older than average, routine-only vaccination would leave a critical vaccine coverage gap between WLHIV and all women for up to 80 years –sustaining higher HPV prevalence and CC incidence for WLHIV. While catch-up vaccination for AGYW-LHIV would immediately boost coverage in those <30 years, before most acquire HPV, it could take up to 70 years to maximize its impact (relative to routine-only vaccination) on CC incidence in WLHIV aged 30+. Instead, vaccinating WLHIV 15+ would immediately close the coverage gap and advance annual CC incidence reductions, particularly among those aged 50+, whose maximum reduction would occur 25 years sooner and be 19%-points higher than when vaccinating AGYW-LHIV.

Optimizing HPV vaccine allocation for WLHIV can inform strategies to reduce CC disparities. While ART scale-up and routine CC screening could reduce persistent HR-HPV infections and CC for WLHIV, like others^29^, we showed they are insufficient on their own. Without any vaccination, our models predicted gradual HR-HPV prevalence declines among WLHIV that leave levels high long-term (18-23%), and that CC incidence could rise. South Africa currently uses the bivalent vaccine for routine vaccination, which, according to our models would slowly reduce VT HR-HPV prevalence and CC incidence among WLHIV, taking up to 75 years for each to decline by 50% from baseline. Using the nonavalent vaccine for routine-only vaccination would further impact VT HR-HPV prevalence and accelerate CC incidence declines among WLHIV, each halving compared to baseline within 20 and 52 years, respectively. Despite the benefits of the nonavalent vaccine, the impacts of routine-only vaccination are still more gradual than when catch-up vaccination is implemented. Our qualitative conclusions about the additional benefits of catch-up vaccination for WLHIV remain robust across our range of sensitivity analyses, provided high and long-lasting vaccine-induced immunity for WLHIV.

Despite consensus on its importance, few model-based evaluations of CC control strategies have jointly modelled HPV-HIV^30–33^, a limited number of which examined the impact of HPV vaccination among WLHIV^34–36^. Those that did showed that, while routine vaccination can substantially reduce long-term CC incidence for WLHIV, inequities will persist, with CC rates remaining higher than in all women even with ART scale-up^29, 37^. Using a model calibrated to data up to 2011, Li et al. suggested that routine vaccination would be cost-effective for WLHIV in South Africa, though results were sensitive to assumed HIV-mortality, vaccine efficacy and duration of protection^38^. More recent analyses based on *MicroCOSM-HPV* indicate that catch-up vaccination of WLHIV using the bivalent vaccine could be cost-effective^23^. Our study results align with those of a recent analysis based in KwaZulu-Natal which projected that the disparities in CC incidence between WLHIV and all women would only be reduced by targeted interventions for WLHIV (catch-up vaccination for AGYW-LHIV and enhanced CC screening and treatment)^19, 29^. Still, like our findings, disparities for WLHIV will persist long-term^9, 28^.

The *WHO Africa Regional Immunization Technical Advisory Group* recommends prioritizing immunocompromised individuals^35^. These findings provide timely insights for other countries with high HIV prevalence, as HPV vaccination programs are scaled up. Catch-up vaccination for WLHIV would likely require a modest, short-term investment in additional doses^23^. With increased manufacturing alongside changes to one-dose schedules for routine vaccination and more affordable pricing for GAVI-supported countries, it may be possible for countries to secure the supply needed^34–36^. Furthermore, while delivery and reach could be challenging, there is opportunity to integrate HPV vaccination with existing HIV services, with some clinics across Africa already doing so for AGYW-LHIV eligible for the routine program^15, 39^.

Our analyses have some limitations. Firstly, our estimates are based on projections of the HIV and HPV epidemics in South Africa, which could be affected by future demographic and epidemiological trends. To minimize the potential impact of demographic shifts, our overall estimates were age-standardized to the South African UN Population Projections up to 2100^20^. However, future intervention levels –particularly HPV or cervical screening– may impact our results. Our projections are based on 2019 cervical screening coverage and frequencies, which, if increased, may overestimate the impact of catch-up vaccination for WLHIV on CC incidence (i.e., fewer WLHIV may progress to CC if pre-cancerous lesions are detected earlier). Secondly, both models have some structural limitations. For example, the deterministic model aged individuals using exponentially distributed rates, resulting in faster transitions through age compartments. The stochastic nature of the individual-based model resulted in highly variable estimates of age-stratified CC incidence, especially for younger age cohorts. Third, there are still uncertainties around the synergistic risks of HPV and HIV acquisition, and HPV and CC disease progression for WLHIV. To reflect this, we present results from two independently developed models with differences in underlying assumptions about disease progression and parameterization, which were each calibrated under Bayesian frameworks. Finally, *Det_HPV-HIV* begins routine vaccination in 2020, despite South Africa having implemented bivalent vaccination in 2014 and reaching 75% and 61% 1- and 2-dose coverage, respectively, among 15-year-old girls in 2020^40^. Given the slow HPV prevalence decline for WLHIV under bivalent vaccination alone, the six year difference should minimally affect our findings.

To our knowledge, this work is the first to isolate the impact of catch-up vaccination for WLHIV considering both HPV and CC outcomes among WLHIV overall and by age. Further, we offer an understanding of routine vaccination among all women and WLHIV using models that explicitly capture key HIV-HPV interactions and related interventions –an important limitation of prior evaluations^31^. We also incorporate the effect of ART scale-up, providing a realistic representation of evolving HIV care. By exploring a wide range of scenarios for cohort coverage, age eligibility, and vaccine characteristics, we offer a comprehensive overview of how program design could maximize the impact of catch-up vaccination for WLHIV^39^.

## Conclusions

Addressing the unique challenges faced by WLHIV in HPV prevention and control is essential for equitably achieving CC elimination goals. Catch-up HPV vaccination for WLHIV can help reduce their CC burden, with benefits emerging earliest if offered to WLHIV of all ages. Prioritizing nonavalent vaccines and achieving high coverage in WLHIV are important for maximizing benefits.

## Data Availability

No data were produced in the present study.

## Prior posting and presentation

This work is the sole product of the authors and has never been submitted for publication.

## Authors Contributions

CMD, MMR, CvS, MB, M-CB, and MM-G contributed to the study conception and design. MMR, CvS, and M-CB contributed to model development, parameterization, and calibration. Analyses of model outputs were performed by CMD, with support from MMR, CvS, M-CB, and MM-G. The manuscript was drafted by CMD. All authors contributed to the interpretation of results and reviewed the manuscript for important intellectual content. Overall supervision for this project was provided by MM-G and M-CB. All authors approved the final manuscript.

