## Supplementary Materials for "Potential impact of catch-up HPV vaccination on HPV prevalence and cervical cancer incidence among women living with HIV in South Africa: results from two mathematical models"

Carla M Doyle^1^, Minttu M Rönn^2*^, Cari van Schalkwyk^3*^, Marc Brisson^4,5^, Nirali Soni^6^, Marie-Claude Boily^7^, Mathieu Maheu-Giroux^1§^

^*^Contributed equally

^§^Corresponding Author

**Author Affiliations:**

^1^Department of Epidemiology and Biostatistics, School of Population and Global Health, McGill University, Montréal, QC, Canada

^2^Department of Global Health and Population, Harvard T.H. Chan School of Public Health, Boston, MA, USA

^3^The South African Centre for Epidemiological Modelling and Analysis, Stellenbosch University, Stellenbosch, South Africa

^4^Centre de recherche du CHU de Québec, Québec, Canada

^5^Département de médecine sociale et préventive, Université Laval, Québec, Canada

^6^Centre for Health Informatics, Computing, and Statistics, Lancaster University Medical School, Lancaster, United Kingdom

^7^MRC Centre for Global Infectious Disease Analysis, School of Public Health, Imperial College London, London, UK

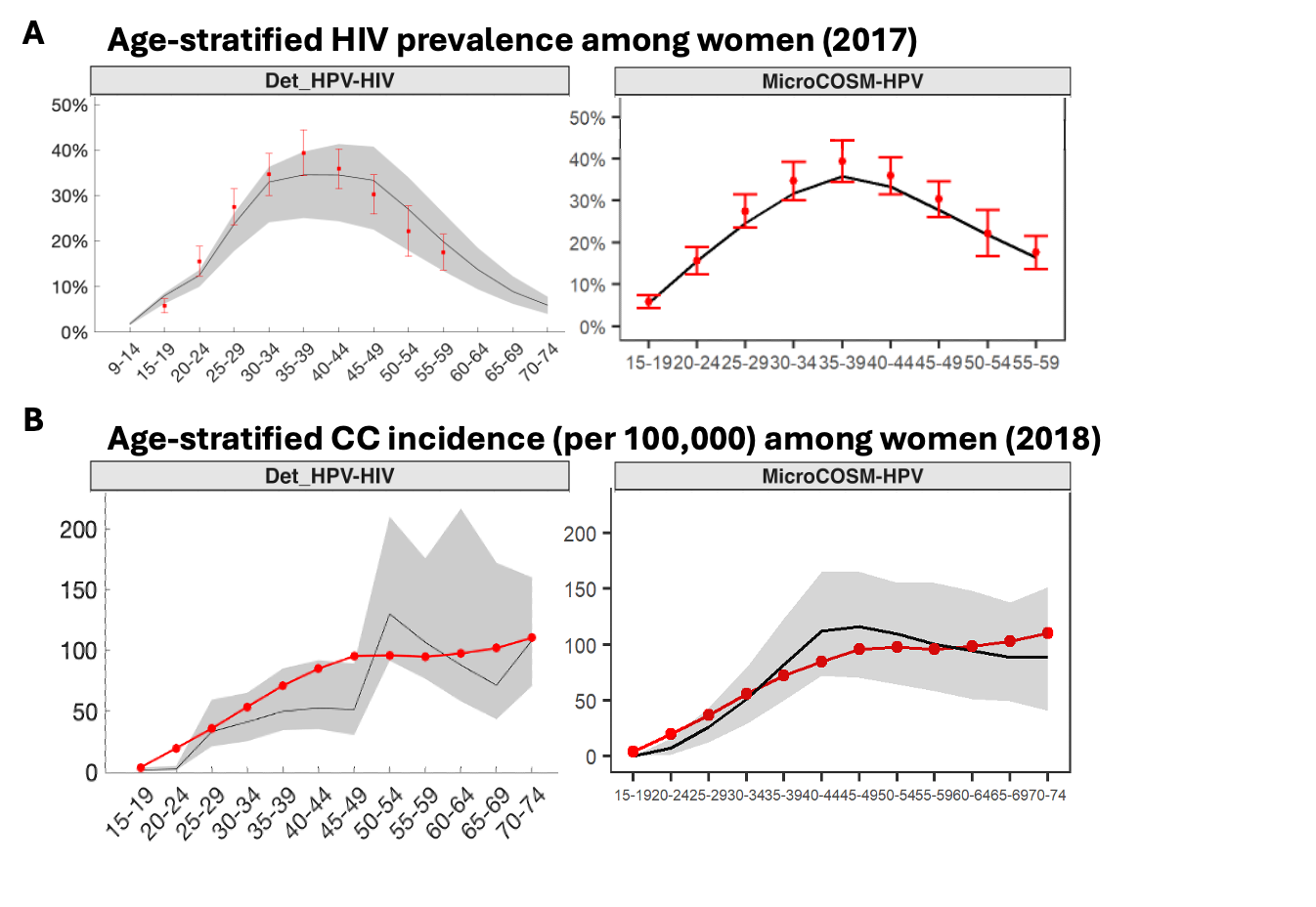

**Figure S1. Select model calibration results reproduced from Boily et al**^1^**.** The age-stratified HIV prevalence in 2017 (Panel A) and cervical cancer incidence in 2018 (Panel B) among women. Each panel presents the median estimates (grey lines) and 90% credible intervals (grey bands) per model. The red points and error bars show the target data used in calibration. For results of the remaining calibration outcomes, please refer to the supplementary materials of the previous model publication by Boily et al^1^.

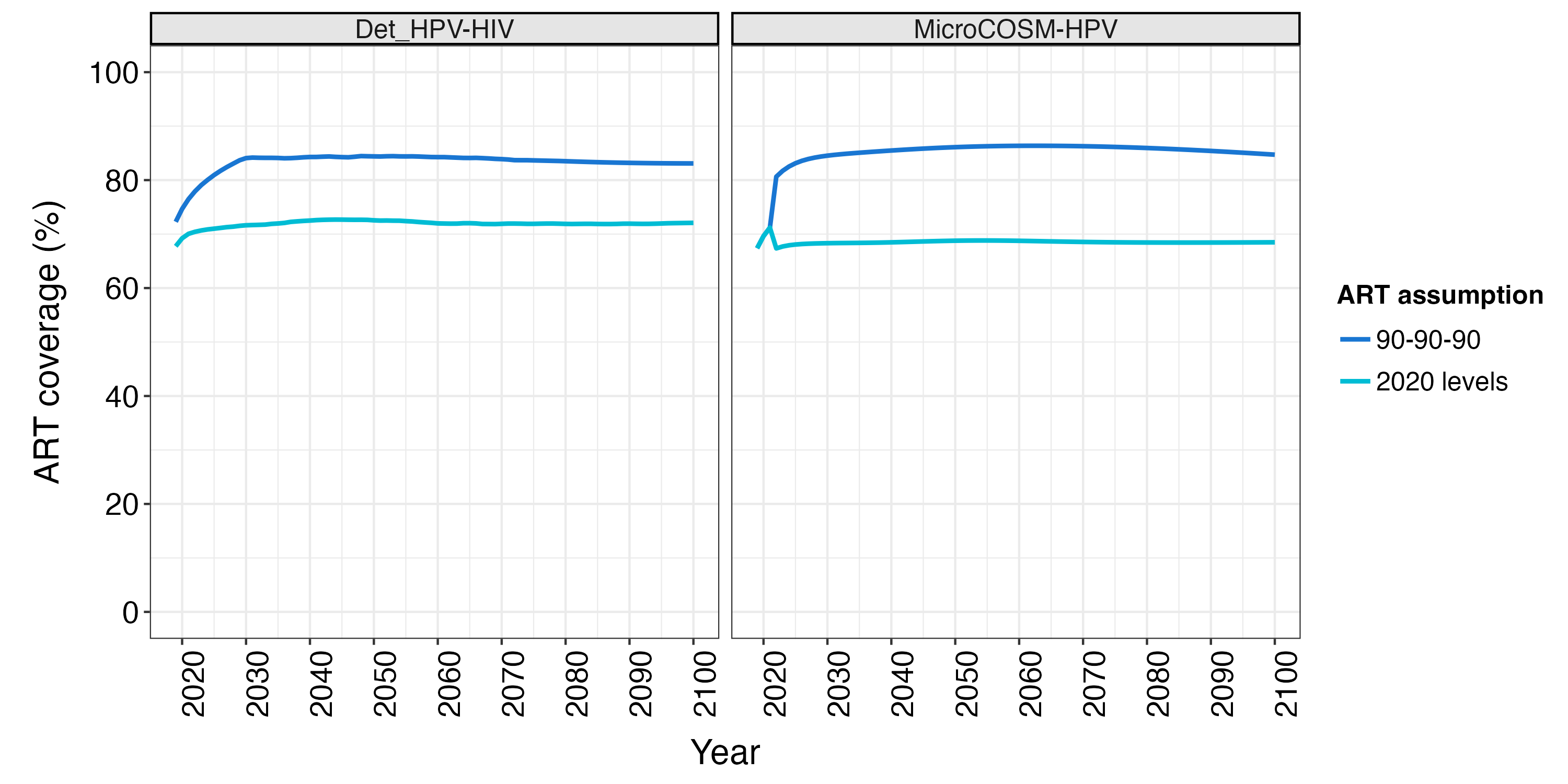

**Figure S2. Antiretroviral therapy (ART) coverage among women living with HIV (WLHIV).** Modelled ART coverage among WLHIV over 2019-2100 under scenarios of routine vaccination in girls aged 9-14 years (90% cohort coverage) with catch-up vaccination for WLHIV aged 15-24 (AGYW-LHIV), with and without ART scale up. *90-90-90* indicates ART scale up to meet the UNAIDS 90-90-90 targets by 2030. *2020 levels* indicates ART coverage plateaus in 2020. The median estimates per model are presented. The colour indicates ART assumption.

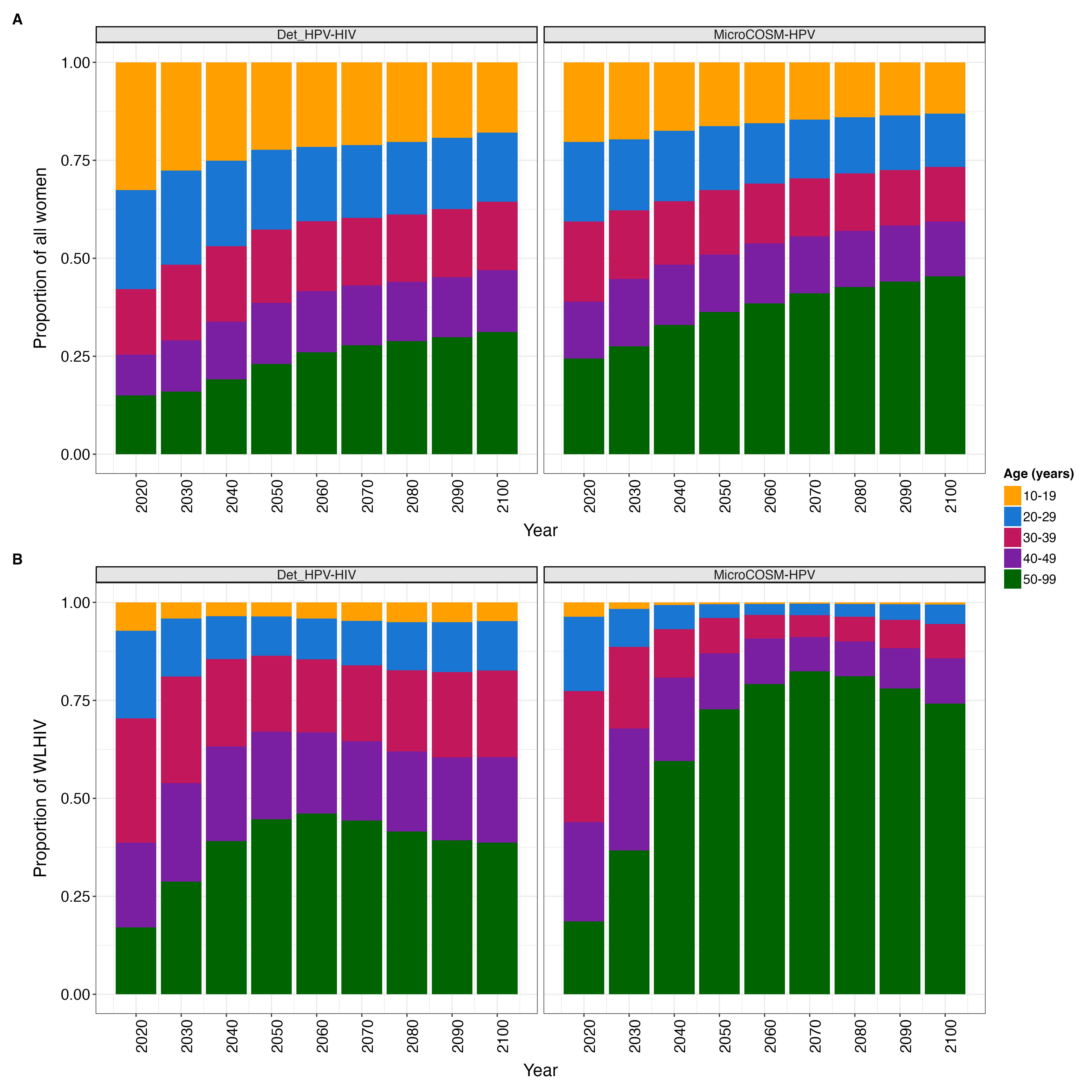

**Figure S3. Model age distribution of all women and women living with HIV (WLHIV) in the absence of routine or catch-up vaccination.** The annual estimated number of women (Panel A) and WLHIV (Panel B) over 2020-2100 when no vaccination is implemented. Each panel presents the median estimates per model. The colour indicates 10-year age categories. The model population in *MicroCOSM-HPV* is scaled up to the population size of South Africa and re-weighted to the population demographics of the South African *Thembisa* model^2^ by age, sex, HIV, and ART status, as it captures the HIV prevention and treatment cascade in more detail.

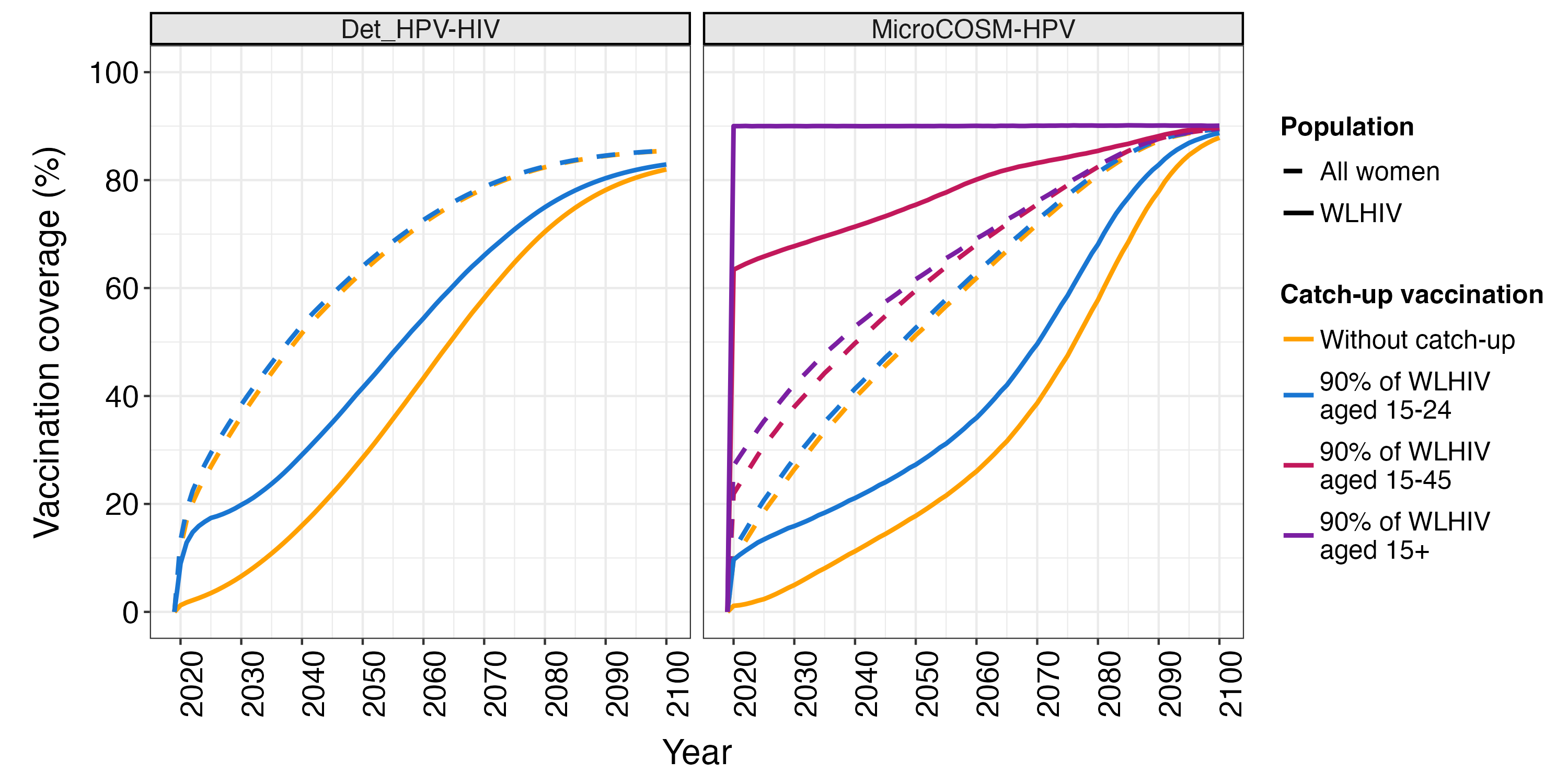

**Figure S4. Vaccination coverage among women living with HIV (WLHIV) and all women under routine and catch-up vaccination.** The modelled vaccination coverage among WLHIV (solid lines) and all women (dashed lines) over 2019-2100 under scenarios of routine vaccination in girls aged 9-14 years (90% cohort coverage), without and with catch-up vaccination for WLHIV aged 15-24 (AGYW-LHIV; 50% and 90% cohort coverage), WLHIV aged 15-45 (90% cohort coverage), and WLHIV aged 15+ (90% cohort coverage). Each panel presents the median estimates per model. The colour indicates the scenario.

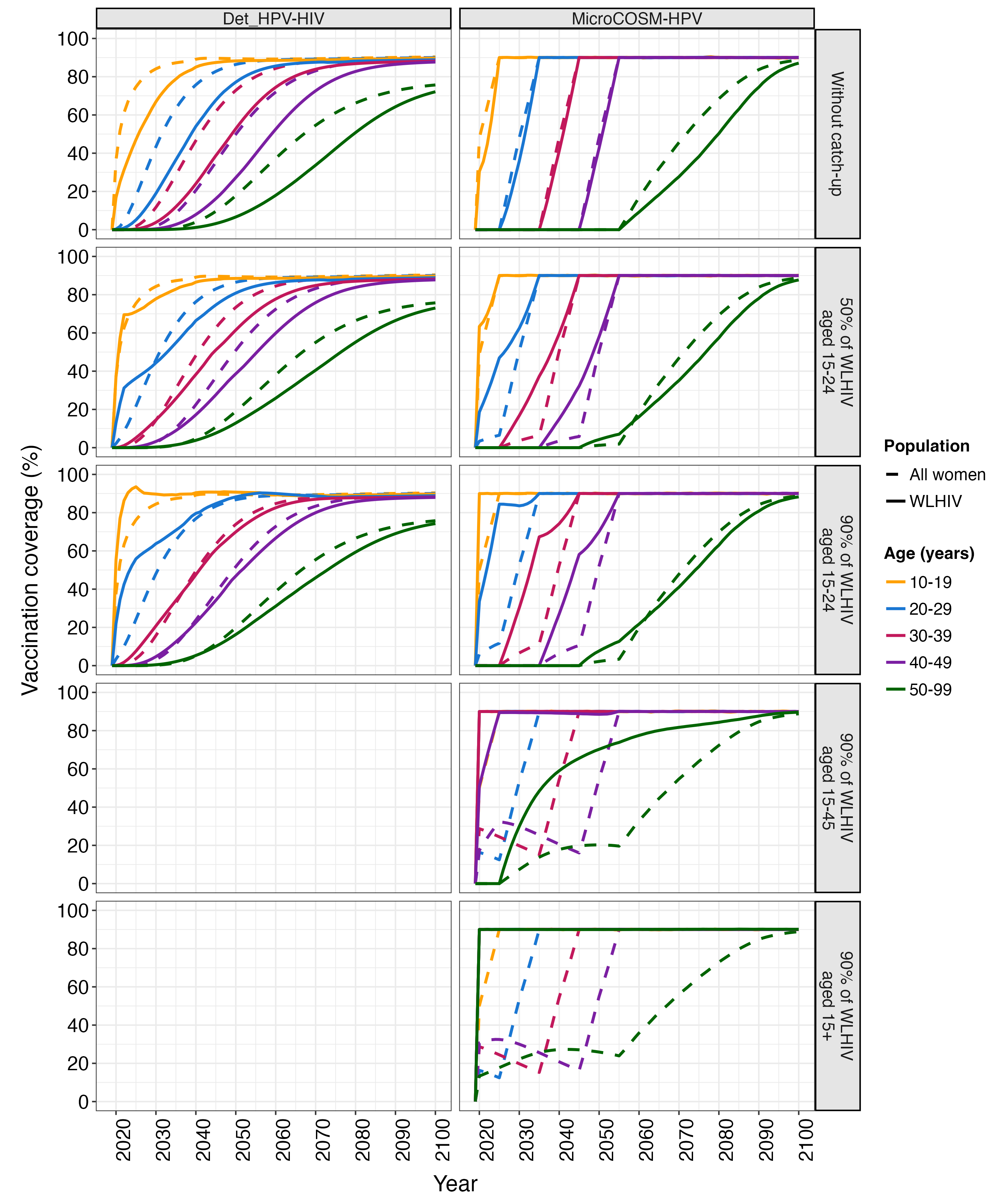

**Figure S5. Vaccination coverage among women living with HIV (WLHIV) and all women by age under routine and catch-up vaccination.** The age-stratified modelled vaccination coverage among WLHIV (solid lines) and all women (dashed lines) over 2019-2100 under scenarios of routine vaccination in girls aged 9-14 years (90% cohort coverage), without and with catch-up vaccination for WLHIV aged 15-24 (AGYW-LHIV; 50% and 90% cohort coverage), WLHIV aged 15-45 (90% cohort coverage), and WLHIV aged 15+ (90% cohort coverage). Each panel presents the median estimates per model. The colour indicates 10-year age categories.

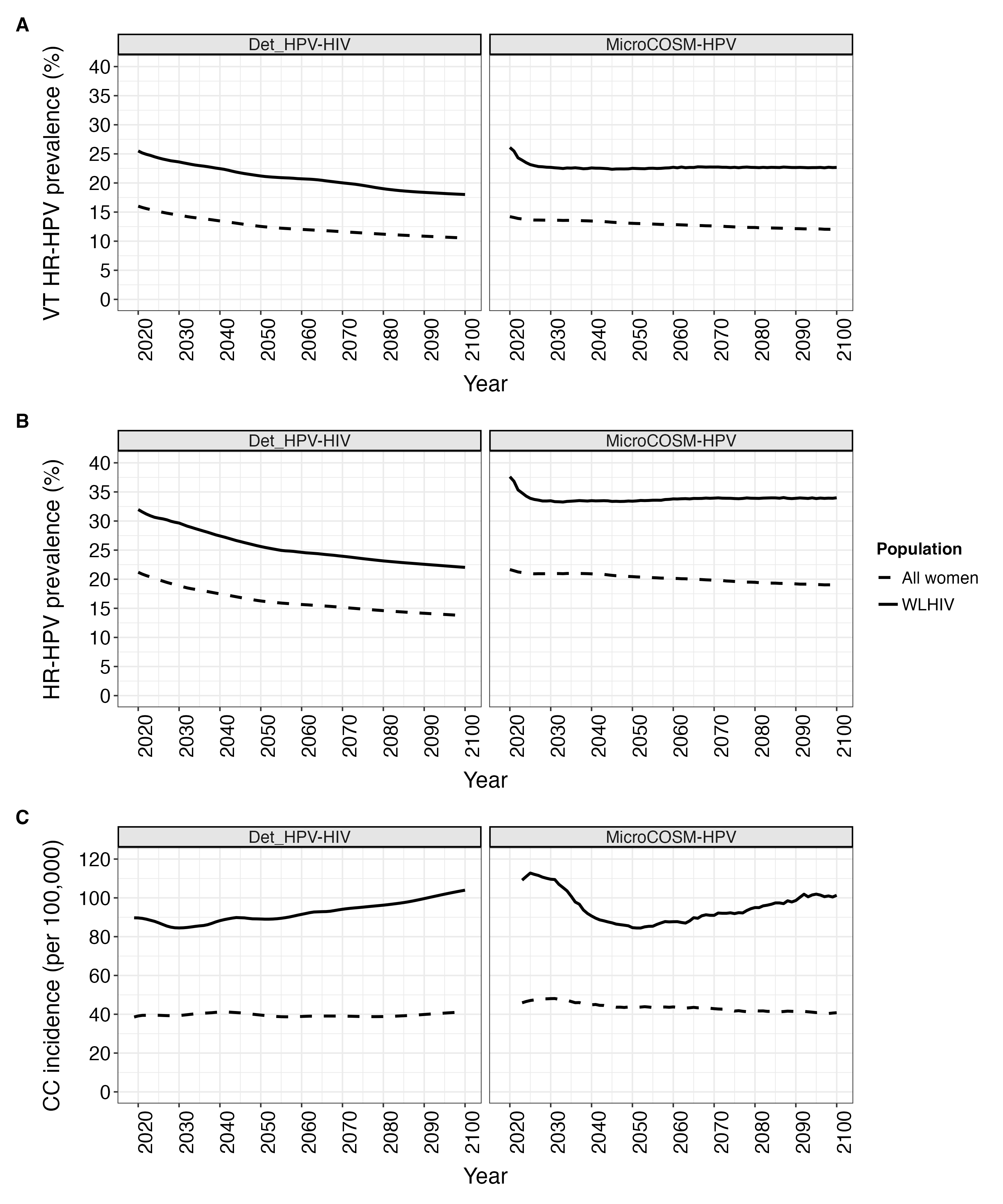

**Figure S6. High-risk HPV (HR-HPV) prevalence and cervical cancer (CC) incidence among women living with HIV (WLHIV) and all women without vaccination.** The predicted annual age-standardized vaccine type (VT; Panel A) and all (Panel B) HR-HPV prevalence and CC incidence (Panel C) among WLHIV (solid lines) and all women (dashed lines) over 2019-2100 in absence of vaccination. Each panel presents the median estimates per model. A 5-year simple moving average was applied to the *MicroCOSM-HPV* CC incidence estimates to smooth stochastic variations.

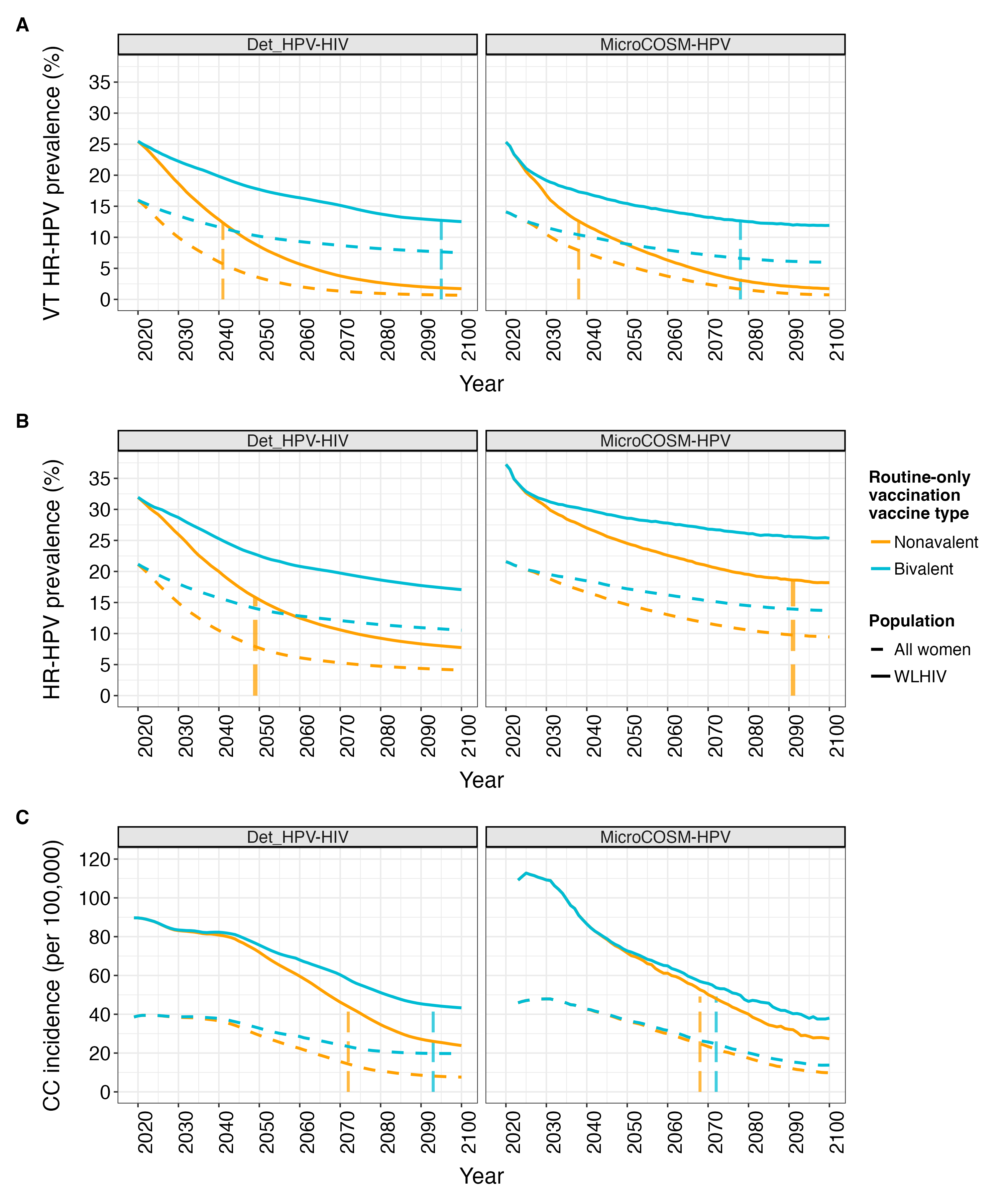

**Figure S7. High-risk HPV (HR-HPV) prevalence and cervical cancer (CC) incidence among women living with HIV (WLHIV) and all women under 90% routine-only vaccination.** The predicted annual age-standardized vaccine type (VT; Panel A) and all (Panel B) HR-HPV prevalence and CC incidence (Panel C) among WLHIV (solid lines) and all women (dashed lines) over 2019-2100 under scenarios of routine vaccination in girls aged 9-14 years (90% cohort coverage) using the bivalent and nonavalent vaccine. Each panel presents the median estimates per model. A 5-year simple moving average was applied to the *MicroCOSM-HPV* incidence estimates to smooth stochastic variations. The colour indicates the scenario. The vertical dashed lines indicate the year when values declined by 50% among WLHIV compared to baseline (2019).

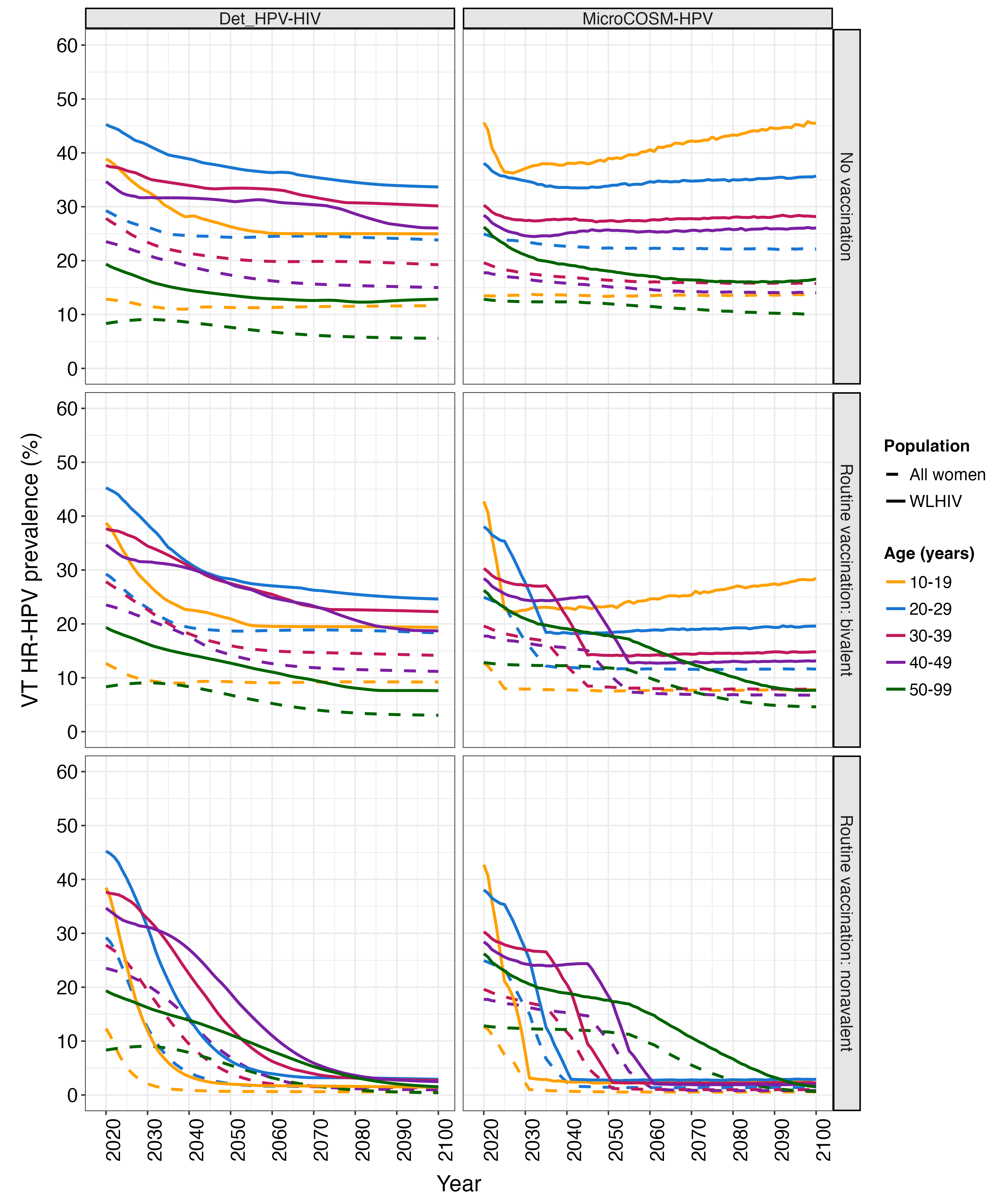

**Figure S8. Impact of routine vaccination on vaccine type (VT) high-risk HPV (HR-HPV) prevalence among women living with HIV (WLHIV) and all women by age: absolute prevalence.** The annual estimated, age-stratified VT HR-HPV prevalence among WLHIV (solid lines) and all women (dashed lines) over 2019-2100 in absence of vaccination and under scenarios of routine vaccination in girls aged 9-14 years (90% cohort coverage) using the bivalent and nonavalent vaccine. Each panel presents the median estimates per model. The colour indicates 10-year age categories.

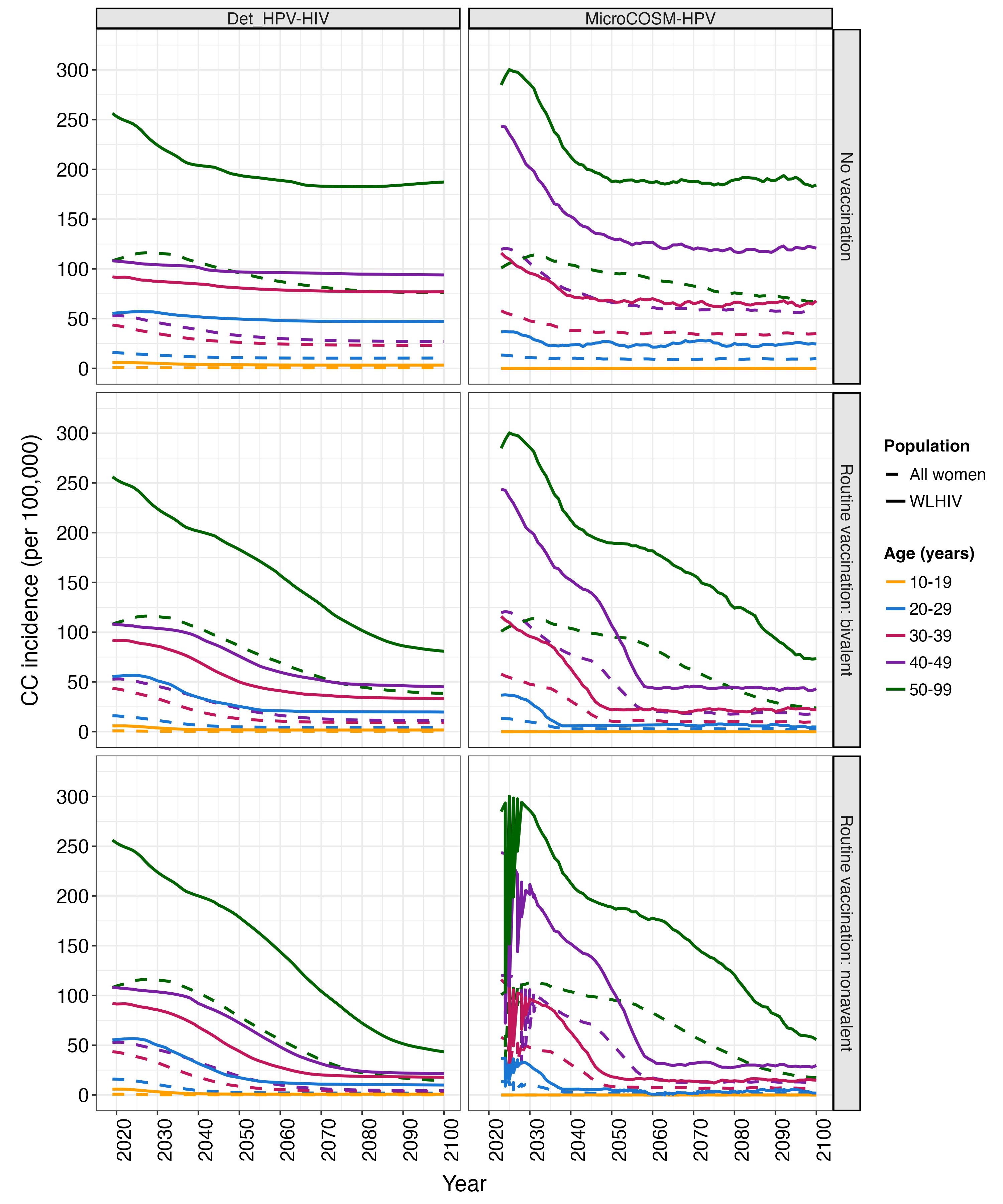

**Figure S9. Impact of routine vaccination on cervical cancer (CC) incidence among women living with HIV (WLHIV) and all women by age: absolute incidence.** The annual estimated, age-stratified CC incidence among WLHIV (solid lines) and all women (dashed lines) over 2019-2100 in absence of vaccination and under scenarios of routine vaccination in girls aged 9-14 years (90% cohort coverage) using the bivalent and nonavalent vaccine. Each panel presents the median estimates per model. The colour indicates 10-year age categories. A 5-year simple moving average was applied to the *MicroCOSM-HPV* incidence estimates to smooth stochastic variations.

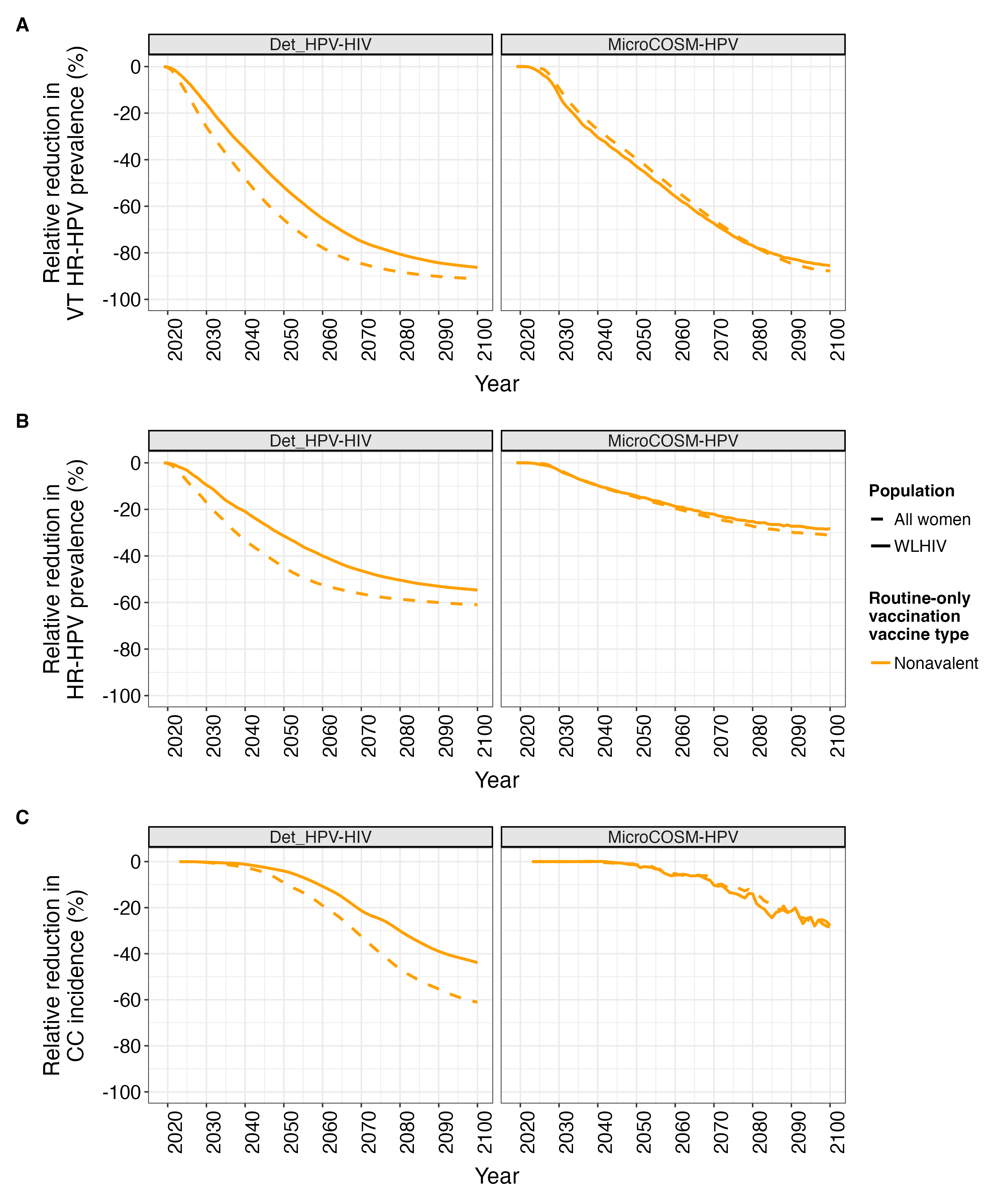

**Figure S10. Impact of nonavalent vs. bivalent vaccination on high-risk HPV (HR-HPV) prevalence and cervical cancer (CC) incidence among women living with HIV (WLHIV) and all women under 90% routine-only vaccination.** The relative reduction in predicted annual age-standardized vaccine type (VT; Panel A) and all (Panel B) HR-HPV prevalence and CC incidence (Panel C) among WLHIV (solid lines) and all women (dashed lines) over 2019-2100 under routine vaccination in girls aged 9-14 years (90% cohort coverage) using the nonavalent vaccine compared to using the bivalent vaccine. Each panel presents the median estimates per model. A 5-year simple moving average was applied to the *MicroCOSM-HPV* incidence reduction estimates to smooth stochastic variations.

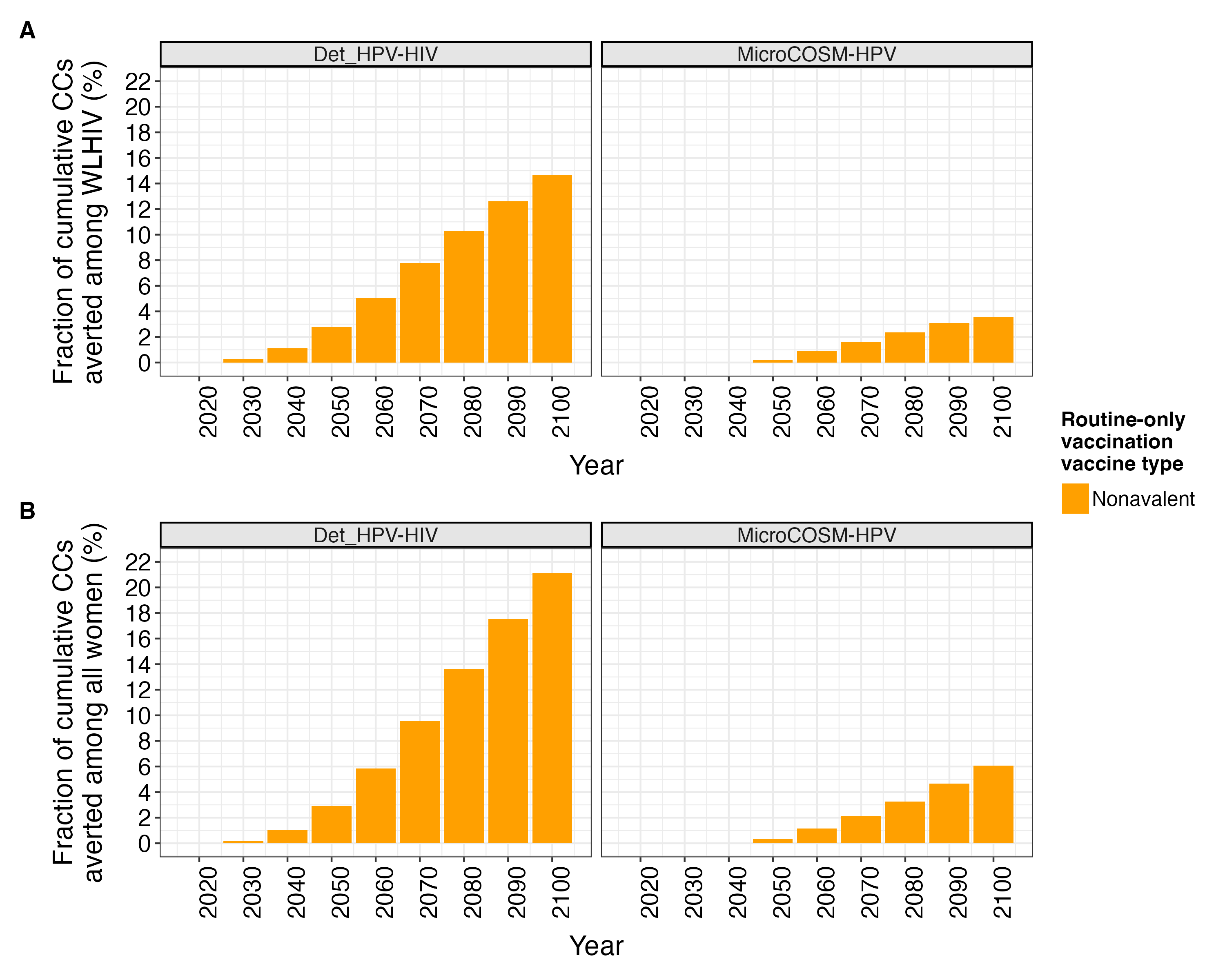

**Figure S11. Cumulative fraction of cervical cancers (CC) averted by nonavalent vs. bivalent vaccination among women living with HIV (WLHIV) and all women under 90% routine-only vaccination.** The annual estimated, age-standardized cumulative fraction of CCs averted among WLHIV (Panel A) and all women (Panel B) over 2020-2100 (at 10-year intervals) under routine vaccination in girls aged 9-14 years (90% cohort coverage) using the nonavalent vaccine compared to using the bivalent vaccine. Each panel presents the median estimates per model.

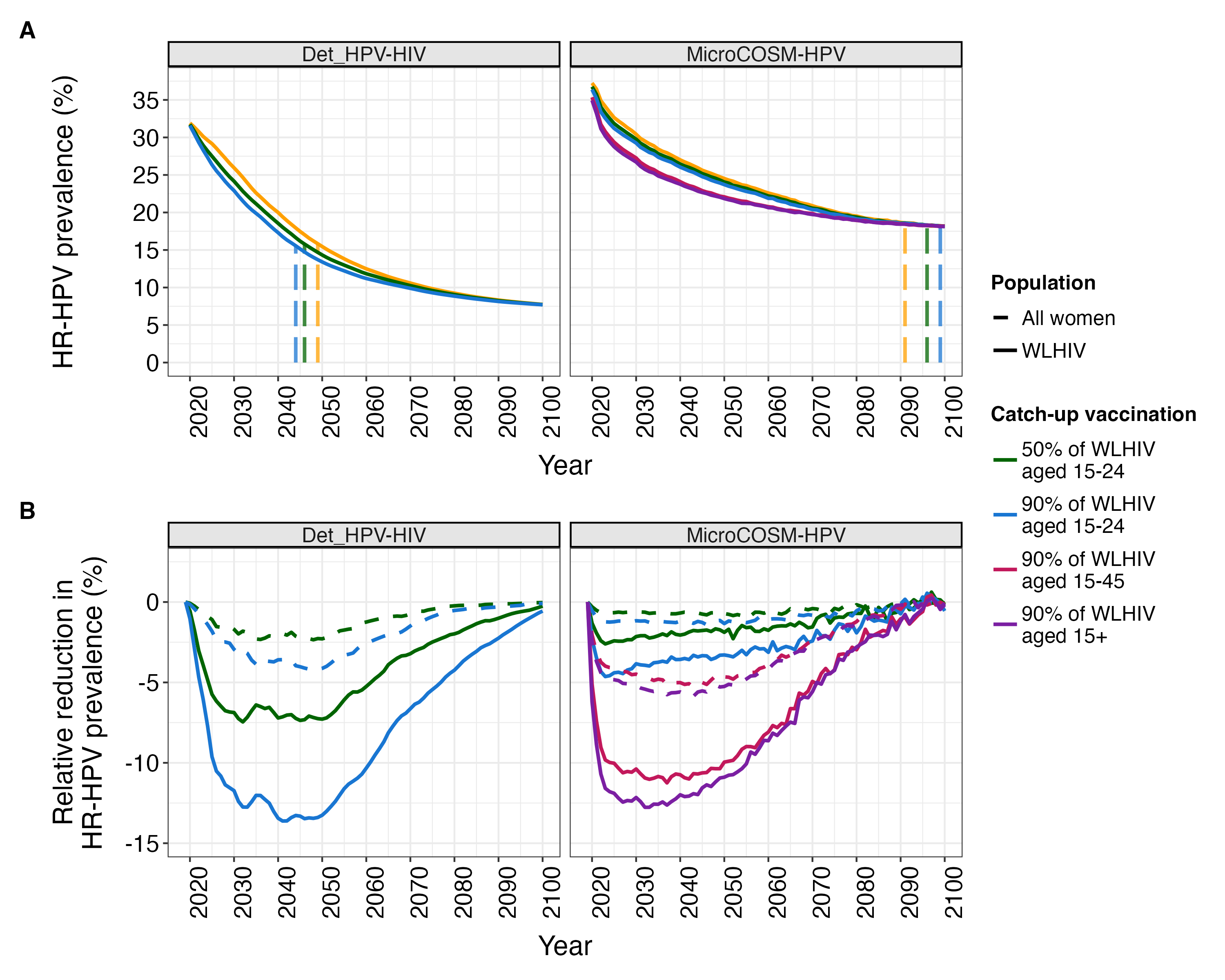

**Figure S12. Impact of catch-up vaccination on high-risk HPV (HR-HPV) prevalence among women living with HIV (WLHIV) and all women.** The annual estimated, age-standardized HR-HPV (all types) prevalence among WLHIV (solid lines) and all women (dashed lines) over 2019-2100 under scenarios of routine vaccination in girls aged 9-14 years (90% cohort coverage) without and with catch-up vaccination for WLHIV aged 15-24 (AGYW-LHIV; 50% and 90% cohort coverage), WLHIV aged 15-45 (90% cohort coverage), and WLHIV aged 15+ (90% cohort coverage). Panel A: absolute prevalence estimates. The vertical dashed lines indicate the year when HR-HPV prevalence declined by 50% among WLHIV compared to baseline (2019). Panel B: relative reduction of catch-up vaccination scenarios compared to routine-only vaccination. Each panel presents the median estimates per model. The colour indicates the scenario.

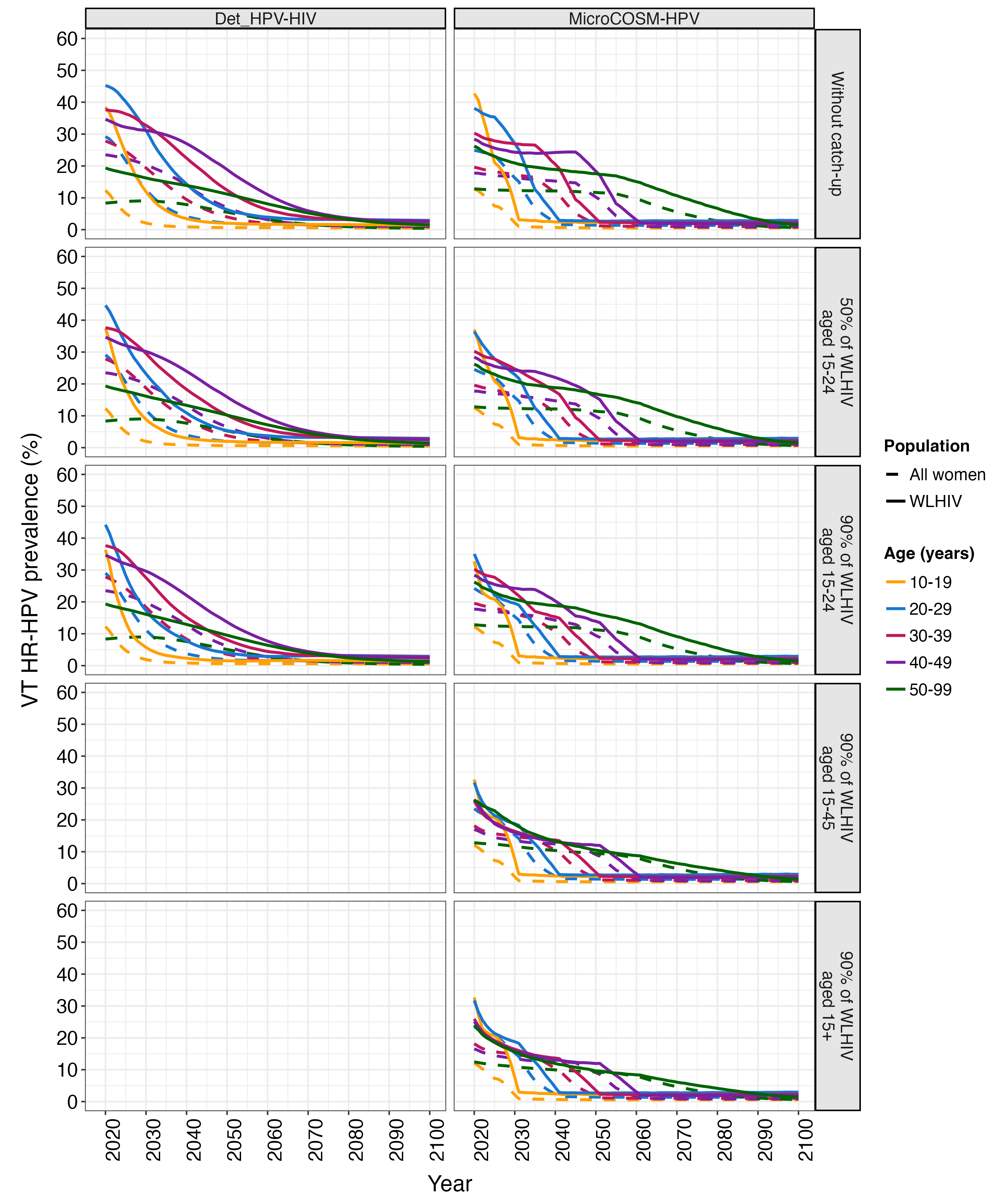

**Figure S13. Impact of catch-up vaccination on vaccine type (VT) high-risk HPV (HR-HPV) prevalence among women living with HIV (WLHIV) and all women by age: absolute prevalence.** The annual estimated, age-stratified VT HR-HPV prevalence among WLHIV (solid lines) and all women (dashed lines) over 2029-2100 under scenarios of routine vaccination in girls aged 9-14 years (90% cohort coverage) without and with catch-up vaccination for WLHIV aged 15-24 (AGYW-LHIV; 50% and 90% cohort coverage), WLHIV aged 15-45 (90% cohort coverage), and WLHIV aged 15+ (90% cohort coverage). Each panel presents the median estimates per model. The colour indicates 10-year age categories.

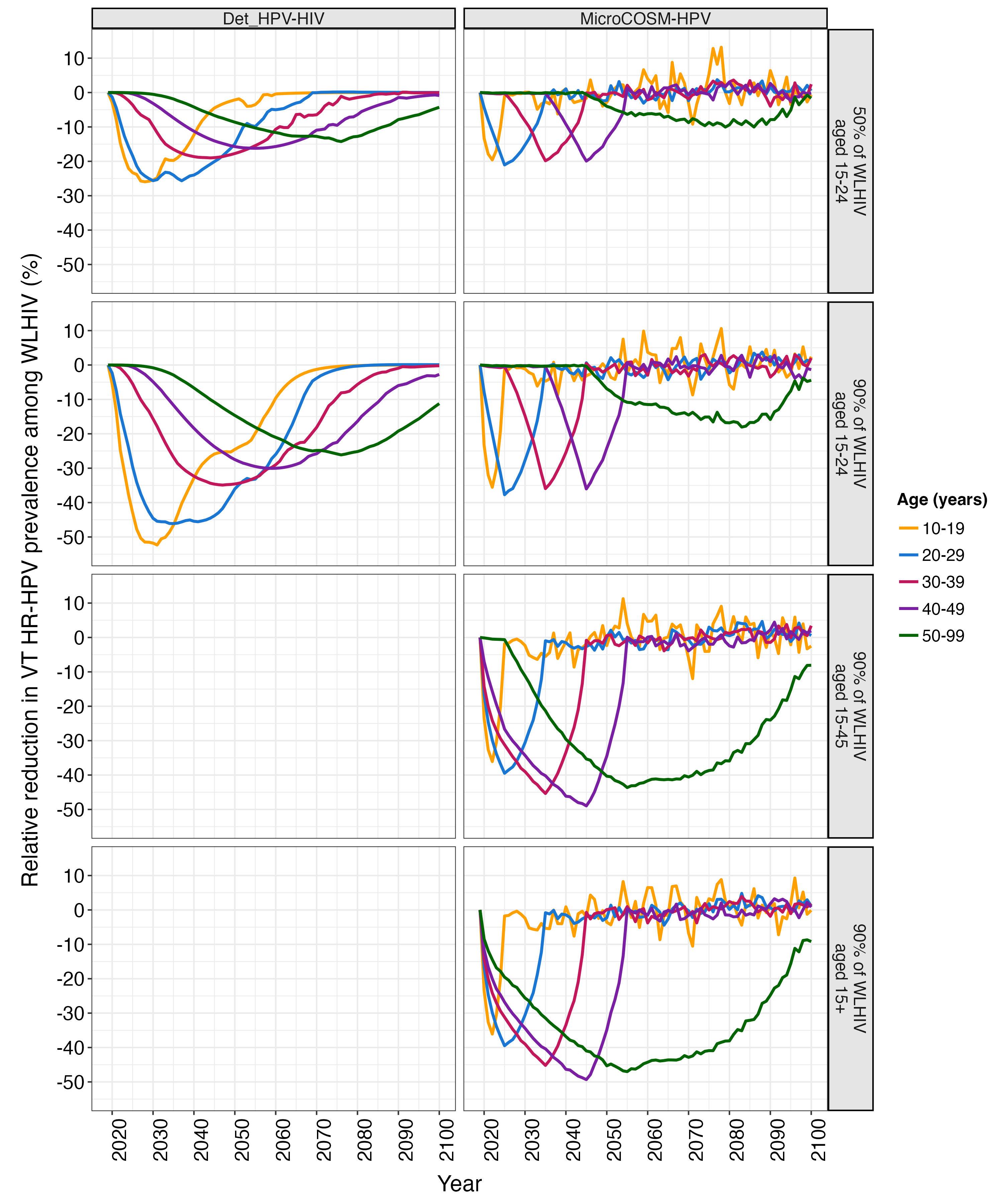

**Figure S14. Impact of catch-up vaccination on vaccine type (VT) high-risk HPV (HR-HPV) prevalence among women living with HIV (WLHIV) by age: relative reductions.** The annual estimated, age-stratified relative reduction in VT HR-HPV prevalence among WLHIV over 2019-2100 under scenarios of routine vaccination in girls aged 9-14 years (90% cohort coverage) with catch-up vaccination for WLHIV aged 15-24 (AGYW-LHIV; 50% and 90% cohort coverage), WLHIV aged 15-45 (90% cohort coverage), and WLHIV aged 15+ (90% cohort coverage) compared to routine-only vaccination. Each panel presents the median estimates per model. The colour indicates 10-year age categories.

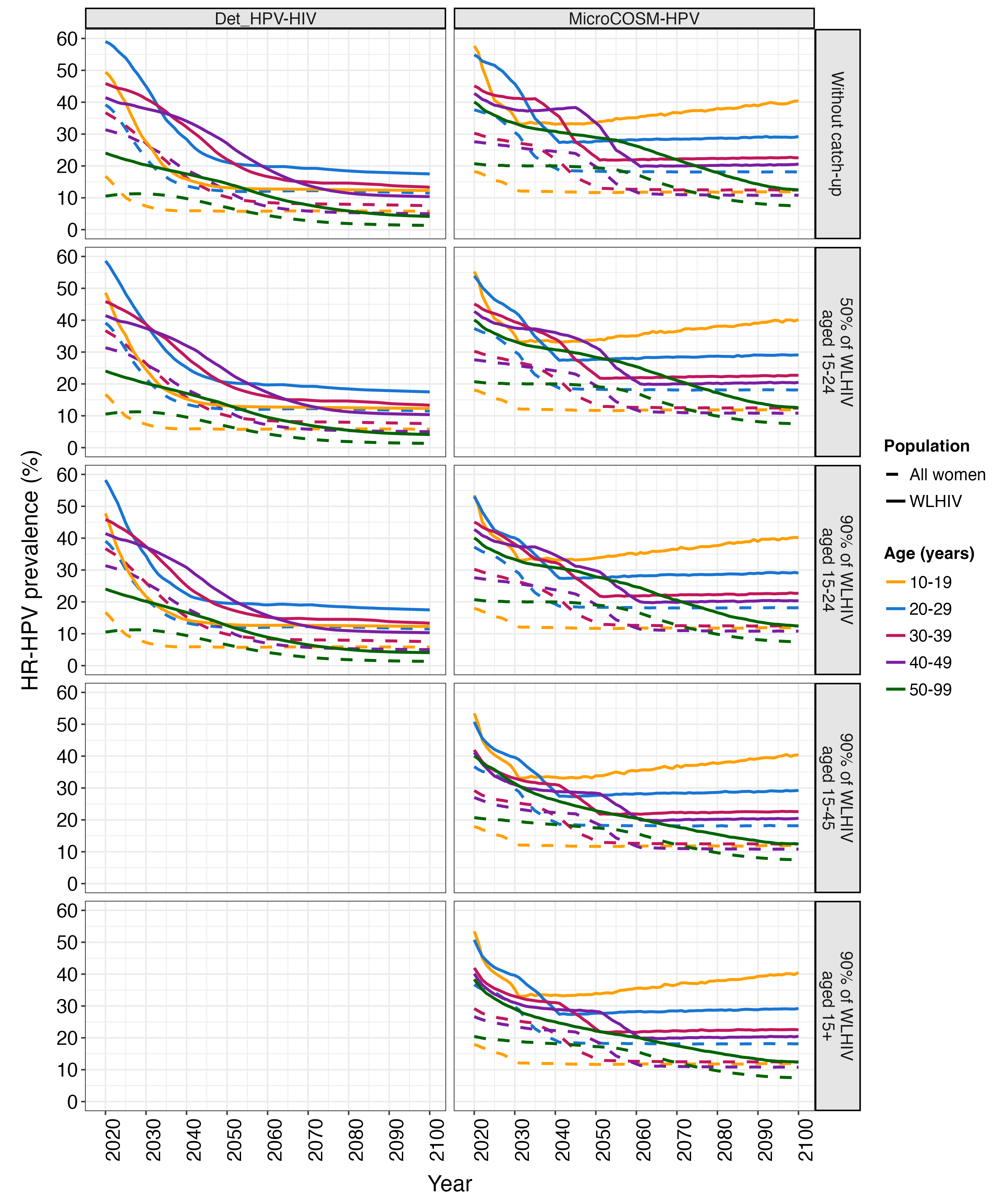

**Figure S15. Impact of catch-up vaccination on high-risk HPV (HR-HPV) prevalence among women living with HIV (WLHIV) and all women by age: absolute prevalence.** The annual estimated, age-stratified HR-HPV (all types) prevalence among WLHIV (solid lines) and all women (dashed lines) over 2019-2100 under scenarios of routine vaccination in girls aged 9-14 years (90% cohort coverage) without and with catch-up vaccination for WLHIV aged 15-24 (AGYW-LHIV; 50% and 90% cohort coverage), WLHIV aged 15-45 (90% cohort coverage), and WLHIV aged 15+ (90% cohort coverage). Each panel presents the median estimates per model. The colour indicates 10-year age categories.

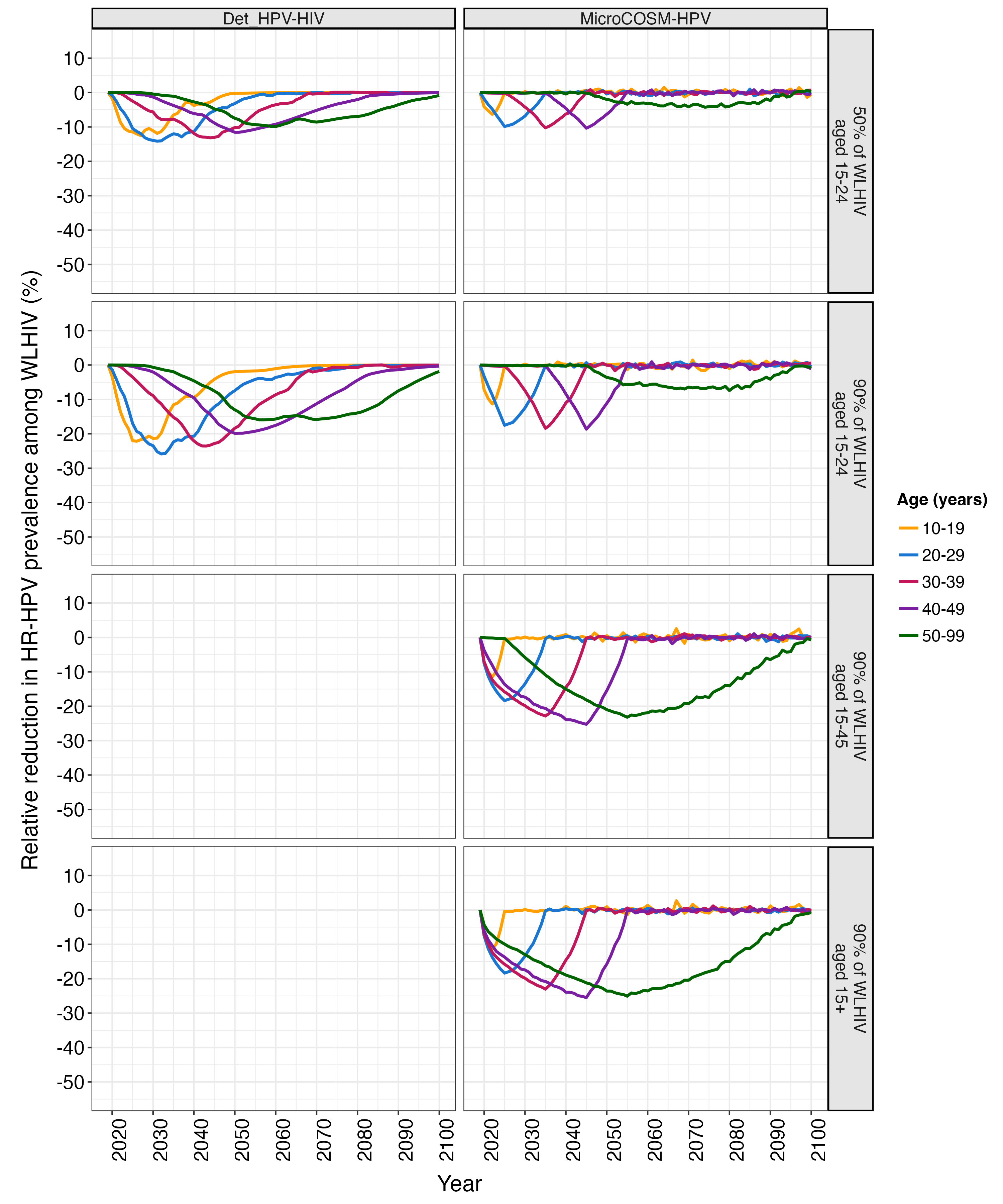

**Figure S16. Impact of catch-up vaccination on high-risk HPV (HR-HPV) prevalence among women living with HIV (WLHIV) by age: relative reductions.** The annual estimated, age-stratified relative reduction in HR-HPV (all types) prevalence among WLHIV over 2019-2100 under scenarios of routine vaccination in girls aged 9-14 years (90% cohort coverage) with catch-up vaccination for WLHIV aged 15-24 (AGYW-LHIV; 50% and 90% cohort coverage), WLHIV aged 15-45 (90% cohort coverage), and WLHIV aged 15+ (90% cohort coverage) compared to routine-only vaccination. Each panel presents the median estimates per model. The colour indicates 10-year age categories.

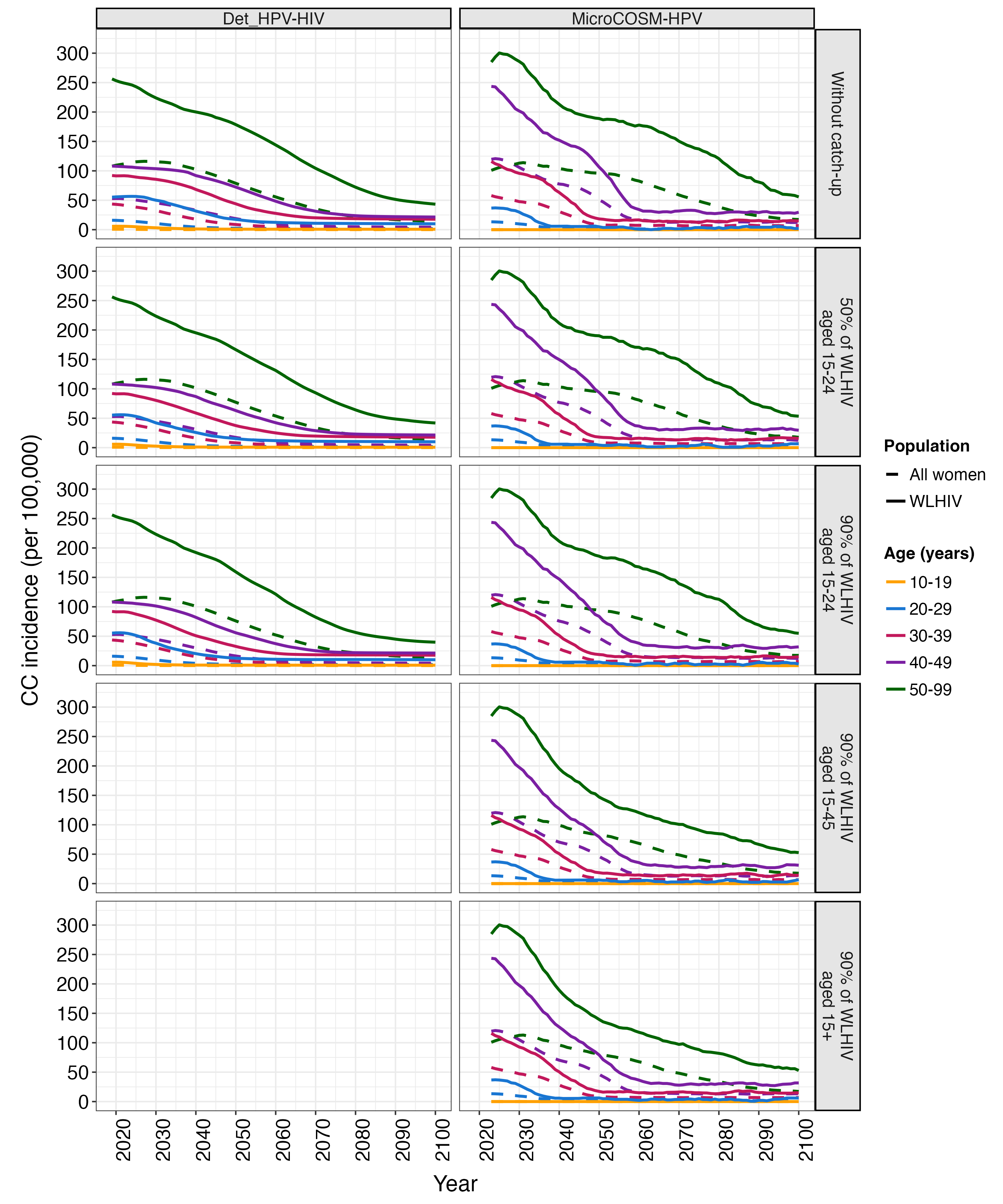

**Figure S17. Impact of catch-up vaccination on cervical cancer (CC) incidence among women living with HIV (WLHIV) and all women by age: absolute incidence.** The annual estimated, age-stratified CC incidence among WLHIV (solid lines) and all women (dashed lines) over 2019-2100 under scenarios of routine vaccination in girls aged 9-14 years (90% cohort coverage) without and with catch-up vaccination for WLHIV aged 15-24 (AGYW-LHIV; 50% and 90% cohort coverage), WLHIV aged 15-45 (90% cohort coverage), and WLHIV aged 15+ (90% cohort coverage). Each panel presents the median estimates per model. The colour indicates 10-year age categories. A 5-year simple moving average was applied to the *MicroCOSM-HPV* estimates to smooth stochastic variations.

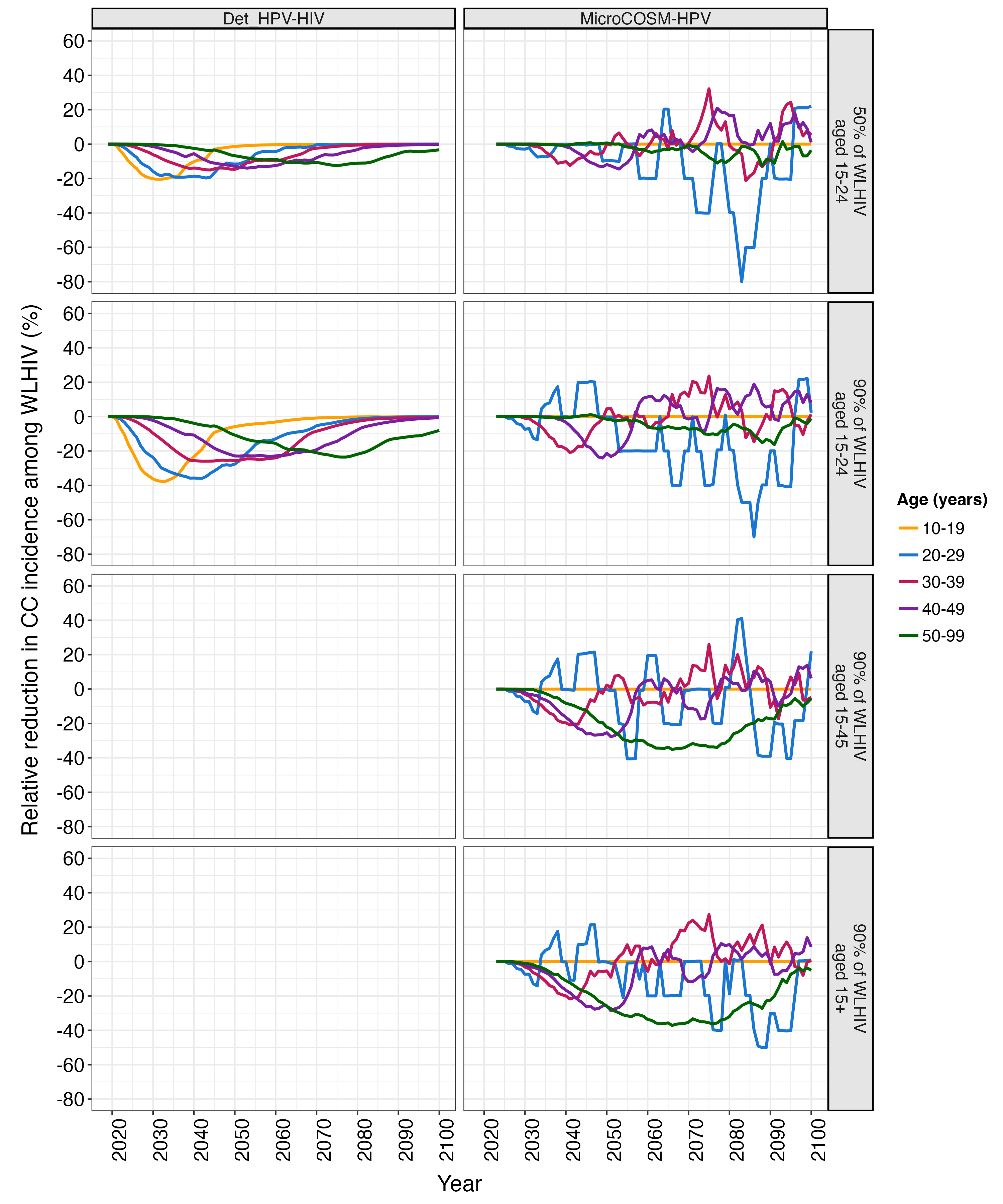

**Figure S18. Impact of catch-up vaccination on cervical cancer (CC) incidence among women living with HIV (WLHIV) by age: relative reductions.** The annual estimated, age-stratified relative reduction in CC incidence among WLHIV over 2019-2100 under scenarios of routine vaccination in girls aged 9-14 years (90% cohort coverage) with catch-up vaccination for WLHIV aged 15-24 (AGYW-LHIV; 50% and 90% cohort coverage), WLHIV aged 15-45 (90% cohort coverage), and WLHIV aged 15+ (90% cohort coverage) compared to routine-only vaccination. Each panel presents the median estimates per model. The colour indicates 10-year age categories. A 5-year simple moving average was applied to the *MicroCOSM-HPV* estimates to smooth stochastic variations.

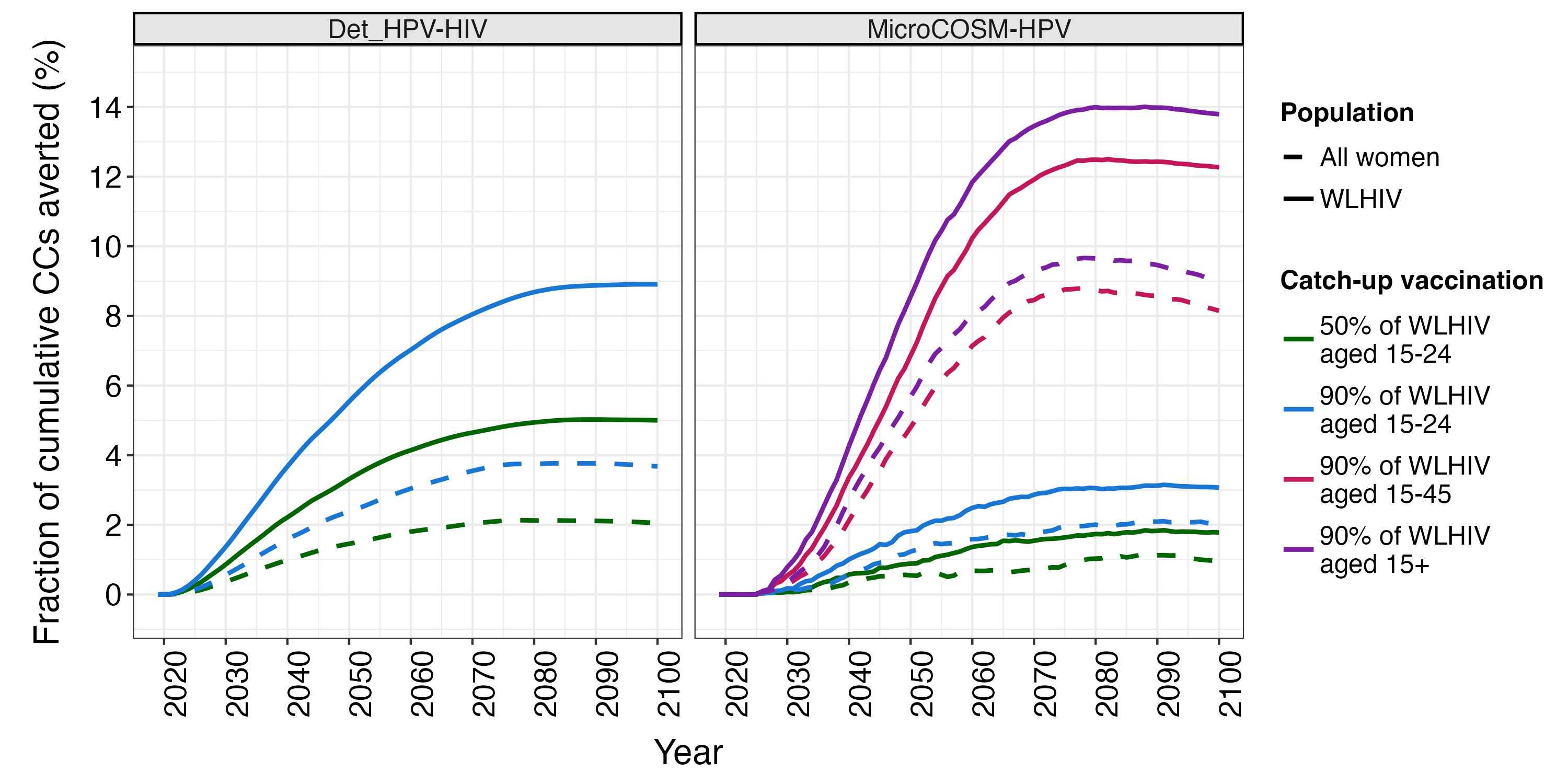

**Figure S19. Cumulative fraction of cervical cancers (CC) averted by catch-up vaccination for women living with HIV (WLHIV) among WLHIV and all women.** The annual estimated, age-standardized cumulative fraction of CCs averted among WLHIV (solid lines) and all women (dashed lines) over 2019-2100 under scenarios of routine vaccination in girls aged 9-14 years (90% cohort coverage) with catch-up vaccination for WLHIV aged 15-24 (AGYW-LHIV), WLHIV aged 15-45, and WLHIV aged 15+ (90% cohort coverage) compared to routine-only vaccination. Each panel presents the median estimates per model. The colour indicates the scenario.

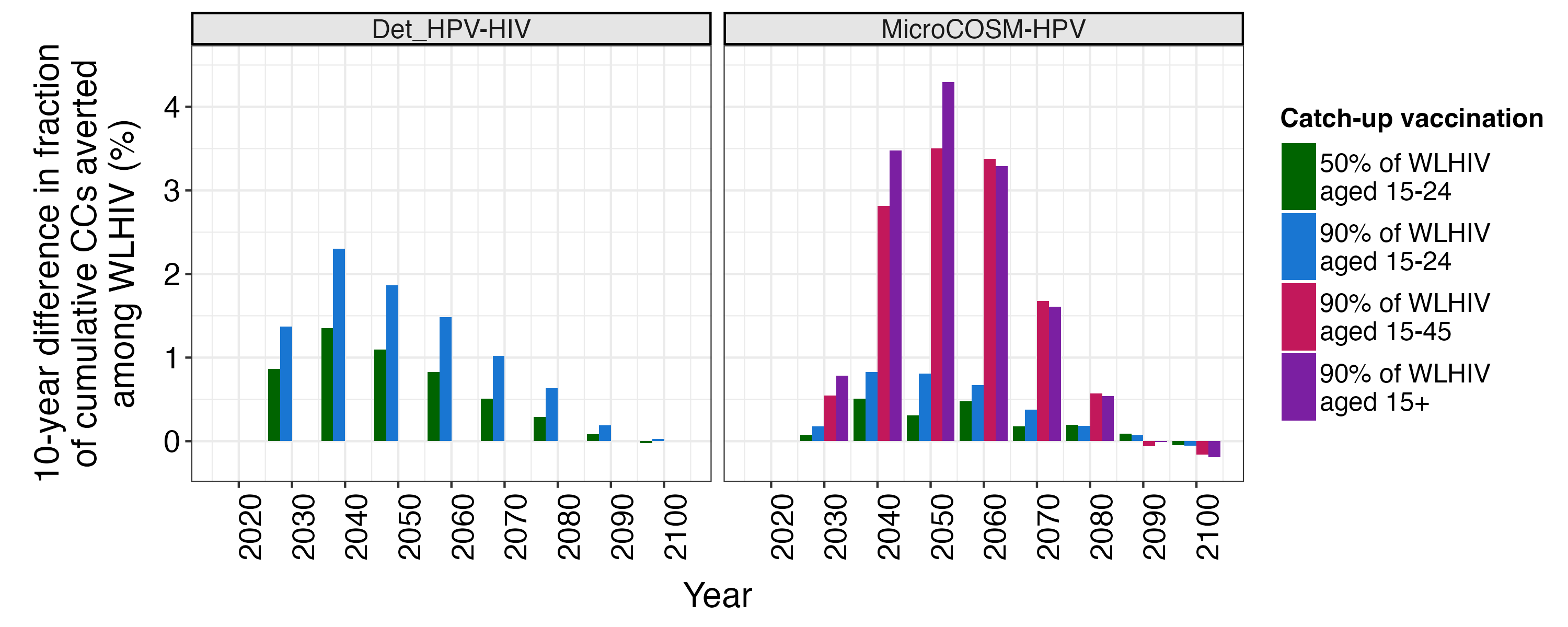

**Figure S20. Ten-year difference in cumulative fraction of cervical cancers (CC) averted by catch-up vaccination for women living with HIV (WLHIV) among WLHIV.** The difference in estimated incremental, age-standardized cumulative fraction of CCs averted at 10-year intervals among WLHIV over 2020-2100 under scenarios of routine vaccination in girls aged 9-14 years (90% cohort coverage) with catch-up vaccination for WLHIV aged 15-24 (AGYW-LHIV; 50% and 90% cohort coverage), WLHIV aged 15-45 (90% cohort coverage), and WLHIV aged 15+ (90% cohort coverage) compared to routine-only vaccination. The 10-year difference in median estimates of the cumulative averted fraction from each model are presented. The colour indicates the scenario.

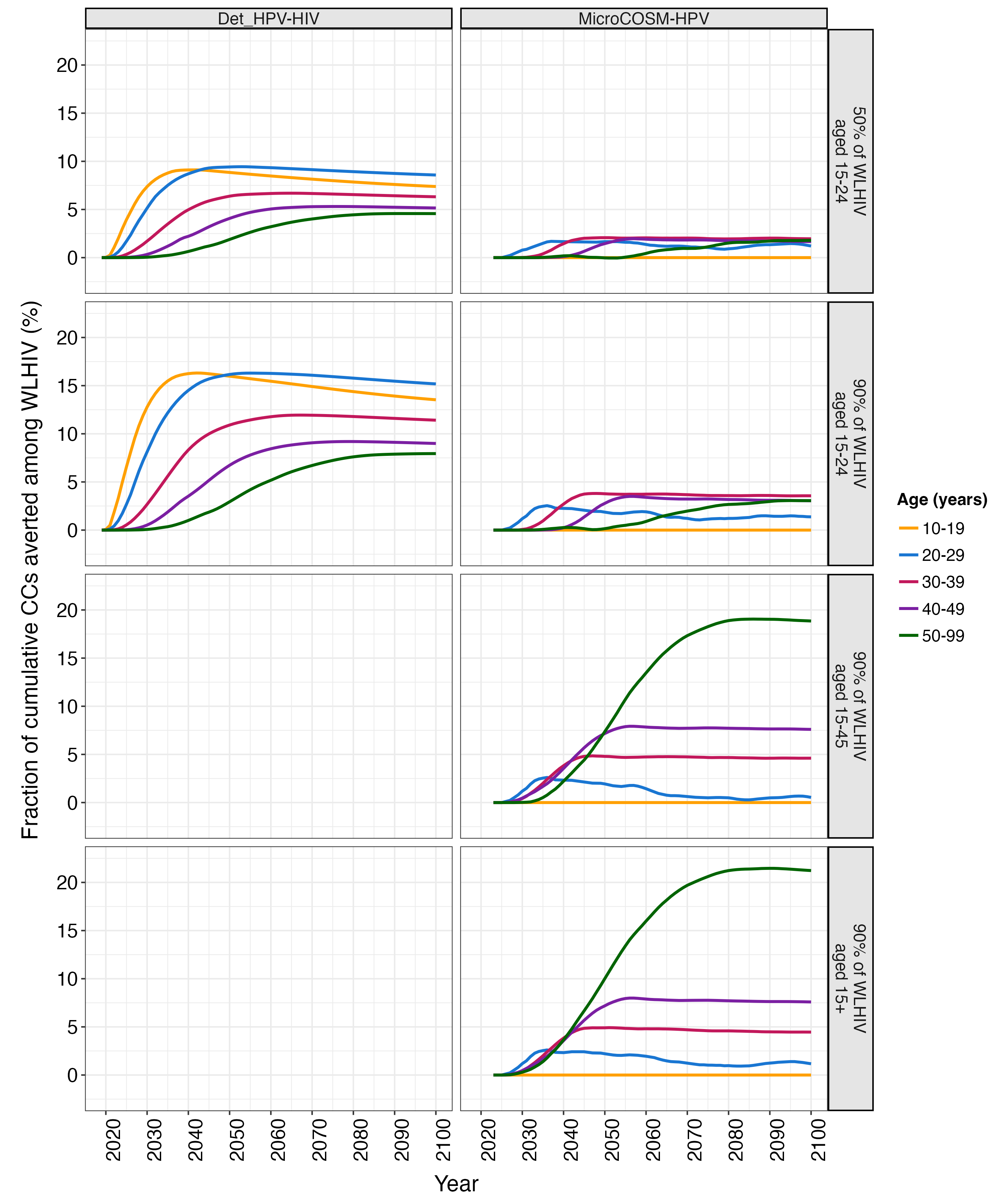

**Figure S21. Age-stratified cumulative fraction of cervical cancers (CC) averted by catch-up vaccination for women living with HIV (WLHIV).** The annual estimated, age-stratified cumulative fraction of CCs averted among WLHIV over 2019-2100 under scenarios of routine vaccination in girls aged 9-14 years (90% cohort coverage) with catch-up vaccination for WLHIV aged 15-24 (AGYW-LHIV; 50% and 90% cohort coverage), WLHIV aged 15-45 (90% cohort coverage), and WLHIV aged 15+ (90% cohort coverage) compared to routine-only vaccination. Each panel presents the median estimates per model. The colour indicates 10-year age categories.

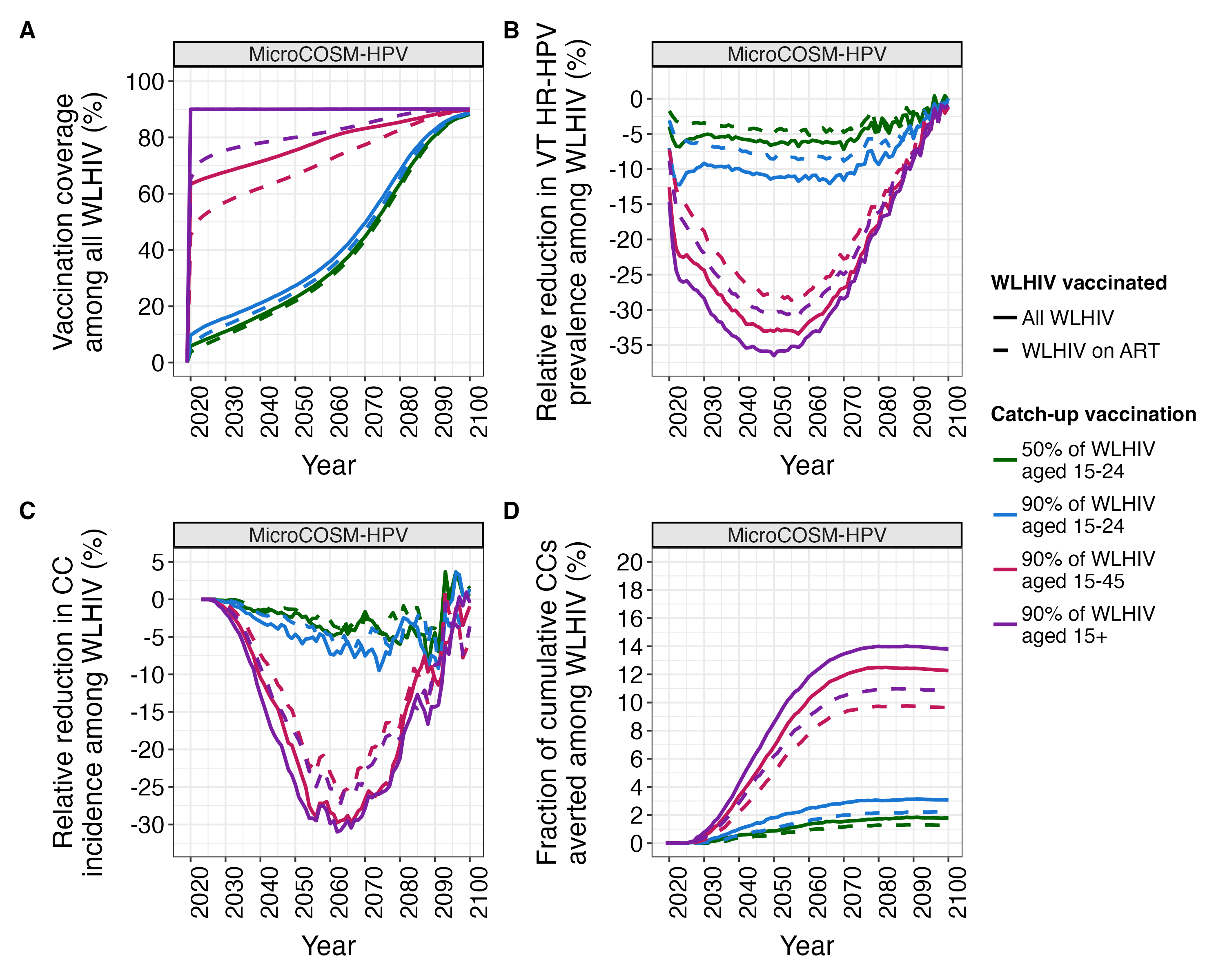

**Figure S22. Impact of catch-up vaccination for women living with HIV (WLHIV) when vaccinating all WLHIV or WLHIV on antiretroviral therapy (ART).** Annual estimated outcomes among WLHIV over 2019-2100 under scenarios of routine vaccination in girls aged 9-14 years (90% cohort coverage) with catch-up vaccination for WLHIV aged 15-24 (AGYW-LHIV), WLHIV aged 15-45, and WLHIV aged 15+ (90% cohort coverage) when modelling catch-up vaccination for all WLHIV (solid lines) or only WLHIV on ART (dashed lines). Panel A: modelled vaccination coverage among all WLHIV. Panels B-D: relative reductions in annual, age-standardized vaccine type (VT) high-risk HPV (HR-HPV) prevalence (Panel B) and cervical cancer (CC) incidence (Panel C), and the annual, age-standardized cumulative fraction of CCs averted (Panel D) by catch-up vaccination scenarios compared to routine-only vaccination. Each panel presents the median estimates per model. The colour indicates the scenario.
